# Alzheimer’s disease protective allele of *Clusterin* modulates neuronal excitability through lipid-droplet-mediated neuron-glia communication

**DOI:** 10.1101/2024.08.14.24312009

**Authors:** Xiaojie Zhao, Yan Li, Siwei Zhang, Ari Sudwarts, Hanwen Zhang, Alena Kozlova, Matthew J. Moulton, Lindsey D. Goodman, Zhiping P. Pang, Alan R. Sanders, Hugo J. Bellen, Gopal Thinakaran, Jubao Duan

**Affiliations:** Center for Psychiatric Genetics, NorthShore University HealthSystem, Evanston, IL 60201, USA; Department of Psychiatry and Behavioral Neuroscience, University of Chicago, Chicago, IL 60637, USA; Department of Bioinformatic, Nanjing Medical University, Nanjing, Jiangsu, 211166, China; Byrd Alzheimer’s Center and Research Institute, University of South Florida, Tampa, FL 33613, USA; Department of Molecular Medicine, Morsani College of Medicine, University of South Florida, Tampa, FL 33160, USA; Jan and Dan Duncan Neurological Research Institute, Texas Children’s Hospital, Houston, TX 77030, USA; Department of Molecular and Human Genetics, Baylor College of Medicine, Houston, TX 77030, USA; Department of Neuroscience and Cell Biology, Child Health Institute of New Jersey, Rutgers Robert Wood Johnson Medical School, New Brunswick, NJ 08901, USA; Department of Neuroscience, Baylor College of Medicine, Houston, TX 77030, USA

**Keywords:** Alzheimer’s disease, Clusterin, protective allele, lipid droplets, neuron excitability, allele-specific open chromatin, genome-wide association study, iPSC, neuron-glia lipid transfer

## Abstract

Genome-wide association studies (GWAS) of Alzheimer’s disease (AD) have identified a plethora of risk loci. However, the disease variants/genes and the underlying mechanisms remain largely unknown. For a strong AD-associated locus near *Clusterin* (*CLU*), we tied an AD protective allele to a role of neuronal CLU in promoting neuron excitability through lipid-mediated neuron-glia communication. We identified a putative causal SNP of *CLU* that impacts neuron-specific chromatin accessibility to transcription-factor(s), with the AD protective allele upregulating neuronal *CLU* and promoting neuron excitability. Transcriptomic analysis and functional studies in induced pluripotent stem cell (iPSC)-derived neurons co-cultured with mouse astrocytes show that neuronal CLU facilitates neuron-to-glia lipid transfer and astrocytic lipid droplet formation coupled with reactive oxygen species (ROS) accumulation. These changes cause astrocytes to uptake less glutamate thereby altering neuron excitability. Our study provides insights into how CLU confers resilience to AD through neuron-glia interactions.

## Introduction

Alzheimer’s disease (AD) is the most common cause of dementia and represents an enormous social and economic burden ^1^. Despite years of research on the accumulation of amyloid beta (Aβ) and Tau lesions, an effective treatment for AD remains out of reach. Recent genome-wide association studies (GWAS) have identified more than 75 reproducible AD risk loci ^2–7^, providing unprecedented opportunities to elucidate novel disease biology that may facilitate the development of more targeted therapeutics. Among the common GWAS-implicated genetic risk factors, the *apolipoprotein E* (*APOE*) locus is by far the strongest genetic risk factor. The protein coding variant *APOE* _ε_*4* (R112/R158; APOE4) allele increases AD risk while the *APOE* _ε_*2* (C112/C158; APOE2) allele decreases AD risk when compared to the most common allele *APOE* _ε_*3* (C112/R158; APOE3) ^8,9^. Accumulation of Aβ and Tau lesions have been central to the postulated pathogenic mechanism underlying the *APOE* risk locus ^8,9^. More recently, APOE has been implicated in novel molecular and cellular mechanisms related to neuron-glia lipid transfer and astrocytic lipid droplet (LD) accumulation ^10,11–14^. However, for most other AD GWAS loci, the causal disease variant/gene, cell type vulnerability, and the underlying molecular/cellular mechanism remain largely unknown.

A top-ranking AD GWAS risk locus falls near the gene *Clusterin (CLU)* ^2–7^. Like APOE, CLU (also known as APOJ) is an abundant apolipoprotein in the brain ^15,16^. As with APOE, CLU has been associated with Aβ deposition and clearance ^17–19^ and Tau pathology ^20,21^. Unlike APOE that is predominantly expressed in glia, CLU is expressed in both glia and neurons ^22,23^. In the brain, CLU-containing lipoproteins are detected in the cerebrospinal fluid, possibly originating from astrocytes, and they may play a role in cholesterol recycling processes ^16,24–26^. Although the total lipid content of CLU lipoprotein particles is much lower than the lipid content found in ApoE lipoprotein particles, CLU lipoprotein particles in astrocytes show a remarkable phospholipid enrichment over APOE particles ^27^. Plasma CLU has also been shown to confer a protective effect by binding to brain endothelial cells and reducing neuroinflammation in a mouse AD model ^28^. Moreover, a recent study suggests that astrocytic CLU protectively promotes excitatory neurotransmission ^29^. However, the physiological role of neuronal CLU in lipid transfer and metabolism, as well as its relevance to AD, remain largely unexplored.

Whether CLU itself is protective or detrimental to AD pathogenesis is controversial. From a genetic perspective, AD GWAS support the presence of an AD protective allele at the *CLU* locus. The frequently reported AD GWAS risk single nucleotide polymorphisms (SNPs) of *CLU,* rs11136000 and rs11787077, both have an allele frequency of 0.38 in Caucasians and they are less frequently observed in AD individuals than in controls with an AD risk odds ratio (OR) of 0.9. Hence, they are protective alleles for AD ^30–34,2–7^. Indeed, GWAS frequently identify derived alleles that confer protection against diseases like AD, cognitive decline, and age-related macular degeneration (AMD) ^35–37^. For example, *APOE* _ε_*2* is well known to be a protective allele for AD ^38,39^. At a genome-wide level, polygenic resilience scores that aggregate the contributions of common variants that promote resilience to AD can be captured ^40^. To better understand the disease mechanism of the AD risk locus near *CLU*, it is imperative to identify the AD causative variant and investigate the functional consequences of the AD protective allele in disease-relevant cell types.

Similar to most other GWAS loci, the AD-associated locus near *CLU* has multiple equally AD-associated noncoding SNPs, posing a challenge to delineate the causal variant. We have recently developed an allele-specific open chromatin (ASoC) mapping approach that enables a comparison of differential chromatin accessibility of the two alleles of a heterozygous disease risk variant in a single sample. This allows the identification of the most likely functional disease risk variants that affect chromatin accessibility and gene expression ^41,42^. By using the ASoC approach, we systematically identified functional AD-associated variants in various human induced pluripotent stem cell (iPSC)-derived subtypes of neurons and glial cells ^43^ and found that among multiple AD risk variants at the *CLU* locus, only rs1532278 (T/C) was located in an open chromatin region (OCR) in human iPSC-derived neurons. We found that the T allele of the ASoC SNP rs1532278 is a derived allele that is protective from AD. We employed CRISPR-cas9 editing to generate isogenic pairs of iPSCs carrying different alleles of rs1532278, followed by integrative functional genomics, molecular/cellular, and metabolic analyses. We found that the AD protective allele T of rs1532278 specifically elevates neuronal CLU expression, which promotes neuron excitability, neuron-to-glia lipid transfer, and LD accumulation in astrocytes. We also show that the elevated astrocytic LDs and reactive oxygen species (ROS) accumulation may contribute to maintenance of neuron excitability by fine-tuning astrocytic glutamate uptake.

## Results

### AD protective allele of rs1532278 elevates *CLU* expression through enhanced ISL2 binding

To study the mechanism underlying the strong AD risk variants near *CLU*, we employed an approach that we recently developed to identify a putatively functional disease causal variant showing ASoC ^41,42^. We screened multiple nearby genome-wide significant (GWS) SNPs that are in strong linkage disequilibrium (r^2^>0.9) with the GWS AD index risk SNP rs11787077 ^44,45^ (**Fig. 1A and B, Table S1**). Using ATAC-seq (assay for transposase-accessible chromatin with sequencing) in iPSC-derived glutamatergic (iGlut), dopaminergic (iDA), GABAergic (iGABA) neurons, astrocytes (iAst), and microglia (iMG) from 39 donors as well as ASoC, we compared the chromatin accessibilities (quantified as ATAC-seq reads) of the two alleles of each heterozygous SNP ^46^. We found that the variant rs11787077 and 9 other GWS SNPs are in strong linkage disequilibrium (r^2^>0.9), but only rs1532278 (T/C) mapped to a strong ATAC-seq peak (i.e., OCR) and also showed a significant ASoC score in iGlut (false discovery rate, FDR<0.033) and iDA (FDR<0.048) neurons but not in iGABA or iAst **(Fig. 1B and C, Table S1**). Note that rs1532278 also showed ASoC in iMG (FDR<0.031) but the SNP-flanking OCR in iMG is minimal compared to neuronal OCRs (**Fig. 1B**). The minor allele T of rs1532278 is a human-specific allele that is less prevalent in AD cases than in controls in GWAS (OR=0.905, P=3.2 × 1010^−33^) (**Fig. S1A**) ^44^. Hence, this suggests that this allele may be functional and protective against AD and warrants further investigation.

**Figure 1.**
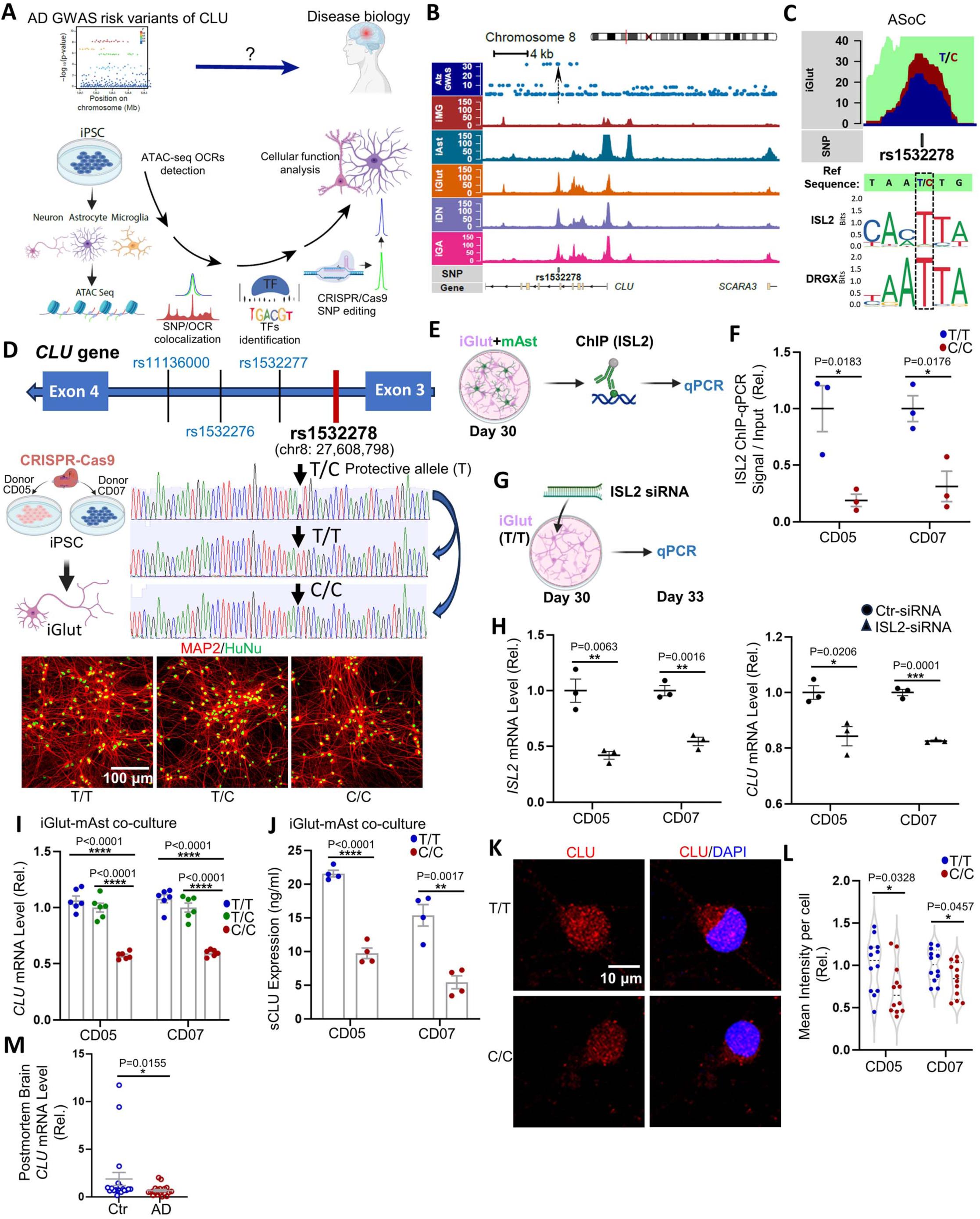
AD risk SNP rs1532278 shows allele-specific open chromatin (ASoC) and *cis*-regulates CLU expression in iGlut. (A) Schematic research design. ATAC-seq was performed to identify OCRs and GWAS risk SNPs that showed ASoC in iPSC Glutaminergic (iGlut), GABAergic (iGABA), and dopaminergic (iDA) neurons, microglia (iMG) and astrocytes (iAst), which is followed by transcription factor (TF) binding prediction to confirm putative functional AD risk SNP, CRIPSR-cas9 SNP editing, and functional assays in cells. (B) ASoC mapping identifies intronic rs1532278 as a putatively functional SNP among several other GWAS risk SNPs equally associated with AD at the *CLU* locus. Note that rs1532278 is the only SNP within the neuronal OCR with a stronger OCR peak in iGlut. (C) T and C alleles of rs1532278 show differential allelic ATAC-seq reads (i.e., ASoC) in iGlut. The bottom panel shows the two most conserved TF binding motifs at the SNP site. (D) Schematic *CLU* gene structure near rs1532278 (upper panel) and a diagram showing CRISPR-Cas9 editing of rs1532278 in two iPSC lines (CD05 and CD07; T/C) to covert T/C lines to isogenic T/T and C/C lines (middle panel). Representative images of iGlut of all three genotypes are also shown (bottom panel); MAP2 and HuNu (human nuclear antigen) staining shows the morphology of iGlut and neuron purity in iGlut-mAst co-cultures. (E-F) ISL2 ChIP-qPCR on day 30 iGlut-mAst co-cultures. n=3 replicates from one clone per line. (G-H) ISL2 siRNA knockdown in day-30 pure iGlut cultures. Samples of 72 hours post-siRNA transfection were used for qPCR. n=3 replicates from one clone per line. (I) Neuronal *CLU* mRNA level in isogenic iGlut-mAst co-cultures of different genotype of rs1532278 (human-specific *CLU* qPCR assay was used). n=6 per group (2-3 clones per line and 2-3 replicates for each clone). (J) Secreted CLU (sCLU) detected by ELISA from the supernatant of iGlut-mAst co-cultures. n=4 per group (2 clones per line, 2 replicates per clone). (K-L) Immunofluorescence staining of CLU in day 23-25 pure iGlut cultures. n=12 coverslips per group of two independent experiments (In total: 2 clones per line and 6 coverslips for each clone; and 5-6 cells per coverslip). (M) *CLU* mRNA level in the gray matter of postmortem brains of patients with AD and controls. Violin plots show data median and interquartile range, all other graphs show data mean±SEM. * *p*<0.05, ** *p*< 0.01, *** *p*< 0.001, and **** *p*< 0.0001. Scale bars are indicated in each image.

We have previously shown that ASoC often reflects differential allelic chromatin accessibility to transcription factor (TF) binding, thereby affecting gene expression ^41,42^. The ASoC SNP rs1532278 is in intron 3 of *CLU* (**Fig. 1D**). By using JASPAR (a database of transcription factor binding profiles) to predict TF binding, we found that the AD protective allele T of rs1532278 is within the TF binding motifs for *ISL2* and *DRGX* (**Fig. 1C**) that are known to be expressed in postmitotic neurons ^47,48^ and in our iGlut (**Fig. S1B**). Interestingly, the more common and ancestral allele C of rs1532278 is predicted to disrupt the binding of these TFs. Moreover, rs1532278 is also in strong linkage disequilibrium with a brain cortex expression quantitative trait loci (eQTL) SNP rs11787077 (r^2^>0.98) whose T allele is associated with increased *CLU* expression ^45^ (**Table S2**). Hence, we postulated that the neuron-specific ASoC SNP rs1532278 is a functional AD risk variant that affects TF binding with the high-affinity allele T likely increasing CLU expression in neurons.

To test this hypothesis, we used CRISPR/Cas9 to edit rs1532278 in two iPSC donor lines (denoted as CD05 and CD07, both homozygous for APOE3) from C/T to T/T or C/C, respectively (**Fig. 1D, Fig. S2A and B**). We then differentiated these iPSC lines into iGlut, a neural subtype that showed more significant ASoC score than iDA (**Fig. 1B and C, Table S1**), and cultured the cells with or without mouse primary astrocytes (mAst) for functional characterizations **(Fig. 1A and D)**. We used mAst instead of human astrocytes to distinguish the source of CLU since it is expressed by both cell types. We first tested whether the SNPs had differential allelic binding to *ISL2* and *DRGX* as predicted (**Fig. 1C**), using chromatin immunoprecipitation qPCR (ChIP-qPCR) (**Fig. 1E**). We compared the TF binding differences within the 120bp SNP-flanking region between the two isogenic pairs in iGlut pure cultures. We found that the two alleles of rs1532278 showed significantly differential chromatin accessibility to *ISL2* (**Fig. 1F**), but not to *DRGX* (**Fig. S1C**), with allele C only retaining 20-30% of *ISL2* binding capacity of allele T (**Fig. 1F**). To further validate whether *ISL2*-binding regulates *CLU* expression, we knocked down (KD) *ISL2* in iGlut neurons carrying the AD risk allele (**Fig. 1G**). We found that ∼50% KD of *ISL2* yielded ∼20% downregulation of *CLU* (**Fig. 1H**). These data indicate that the AD protective allele of rs1532278 has stronger chromatin accessibility to *ISL2* binding, thereby likely increasing *CLU* expression.

We next examined whether rs1532278 could indeed alter *CLU* expression. Culturing iGlut neurons with or without mAst for 30 days, we found that *CLU* mRNA expression was about 40-50% lower in the C/C neurons versus the T/T neurons (**Fig. 1I and S1D**), and the neuronal CLU changes did not affect mouse astrocytic *CLU* expression in the co-culture system (**Fig. S1F**). Using three sets of qPCR primers near the SNP site that cover all major transcript isoforms of *CLU*, we further confirmed that rs1532278 similarly affected these major *CLU* isoforms (**Fig. S1G** and **H**). Since CLU is predominantly secreted (>90%) ^49^, we further measured the levels of the human neuron-secreted CLU protein (sCLU) in the iGlut/mAst co-cultures using an ELISA. Consistent with the allelic effect of rs1532278 on *CLU* mRNA levels, we found that the allele T was associated with higher levels of sCLU in the media (**Fig. 1J and S1E**). Although sCLU is the predominant form, we also detected intracellular CLU in iGlut by immunostaining, with the T allele being associated with higher expression (**Fig. 1K and L**), although to a lesser extent than the mRNA level or sCLU levels (**Fig. 1I and J**). Finally, since rs1532278 is ∼25 kb upstream of *Scavenger Receptor Class A Member 3 (SCARA3)* (**Fig. S1G**), a gene with a plausible role in AD ^50^, we also investigated whether rs1532278 can *cis*-regulate *SCARA3* expression in iGlut. However, we found no significant difference in mRNA expression between the two groups of isogenic neurons with the alleles T and C, suggesting that the functional effect of rs1532278 is restricted to *CLU* (**Fig. S1I**). Altogether, these data show that rs1532278 exerts a *cis*-regulatory effect on *CLU* expression, and the protective T allele is associated with elevated *CLU* expression.

To further support the observed effect of the AD protective allele on neuronal CLU expression, we examined *CLU* expression in post-mortem brains of controls (non-AD) and AD patients. We found significantly higher levels of *CLU* in non-AD controls (**Fig. 1M**). To further delineate the *CLU* levels in different brain cell types in AD patients vs. controls, we mined the single-cell RNA sequencing (scRNA-seq) datasets from postmortem prefrontal cortices ^51,52^. We found that excitatory (i.e., glutamatergic) neurons of non-AD controls showed the most significant elevation of *CLU* (FDR=0), and to a lesser extent in astrocytes (FDR<3.11 × 10^-2^^94^), inhibitory neurons (FDR<5.06 × 10^-2^^10^) and oligodendrocytes (FDR<2.28 × 1010^−25^) of non-AD controls (**Fig. S3A**). The data corroborate our observations of increased expression of *CLU* in iGlut carrying the AD protective allele. However, the AD-associated differential expression of *CLU* in astrocytes and oligodendrocytes in the scRNA-seq dataset seems to be inconsistent with our ATAC-seq data that did not show obvious OCRs or ASoC at the rs1532278 site (**Fig. 1B and Table S1**). We thus tested CLU’s expression in our iAst. We differentiated the CRISPR-engineered isogenic pair of iPSCs carrying T/T or C/C alleles (**Fig.1D**) into iAst ^53^ to compare the allelic effect of rs1532278 on *CLU* expression (**Fig. S3B**). Consistent with our ASoC data, we found no difference in *CLU* expression between these isogenic pairs, confirming that rs1532278 is not a functional SNP in astrocytes (**Fig. S3C**). Hence, elevated *CLU* expression in astrocytes of post-mortem brains of non-AD controls may not be due to the direct effect of the AD risk variant rs1532278 at the *CLU* locus. Alternatively, there may be other AD risk SNPs that may affect *CLU* expression in astrocytes or other cell types. Altogether, these findings show that the AD risk SNP rs1532278 is a neuron-specific functional ASoC SNP at the *CLU* locus, and the protective T allele is associated with higher *CLU* expression in excitatory neurons.

### iGlut carrying the AD protective allele of rs1532278 are more mature and active

A recent study showed that astrocytic CLU can promote excitatory synaptic transmission ^29^. Given our observed neuron-specific effect of the AD protective allele rs1532278 on *CLU* expression in iGlut, we investigated whether the AD protective allele of *CLU* and its associated higher CLU expression have functional effects on neuronal properties. We first immunostained neurons from our iGlut/mAst co-cultures (day-30) using antibodies against MAP2 (microtubular associated protein 2, a neuron-specific cytoskeletal protein), SYP (synaptophysin, a presynaptic marker), and PSD-95 (also known as DLG4, a postsynaptic maker) to analyze the impacts of T/T (protective) versus C/C (risk) alleles on dendritic branching and synapse morphology (**Fig. 2A-E and S4A**). We found that iGlut with the T/T alleles had more dendritic branches (**Fig. 2A and B**). We also found significantly higher SYP puncta density (**Fig. 2C and D**) in T/T iGlut compared with C/C iGlut, but no differences for PSD-95. The immunofluorescence staining results for SYP and PSD-95 were further confirmed by Western blot (WB) analysis (**Fig. 2E and F**). The iGlut monocultures without mAst gave similar results (**Fig. S4B and C**). The neuronal morphometric differences between T/T and C/C iGlut suggest T/T iGlut may exhibit an augmented presynaptic function.

**Figure 2:**
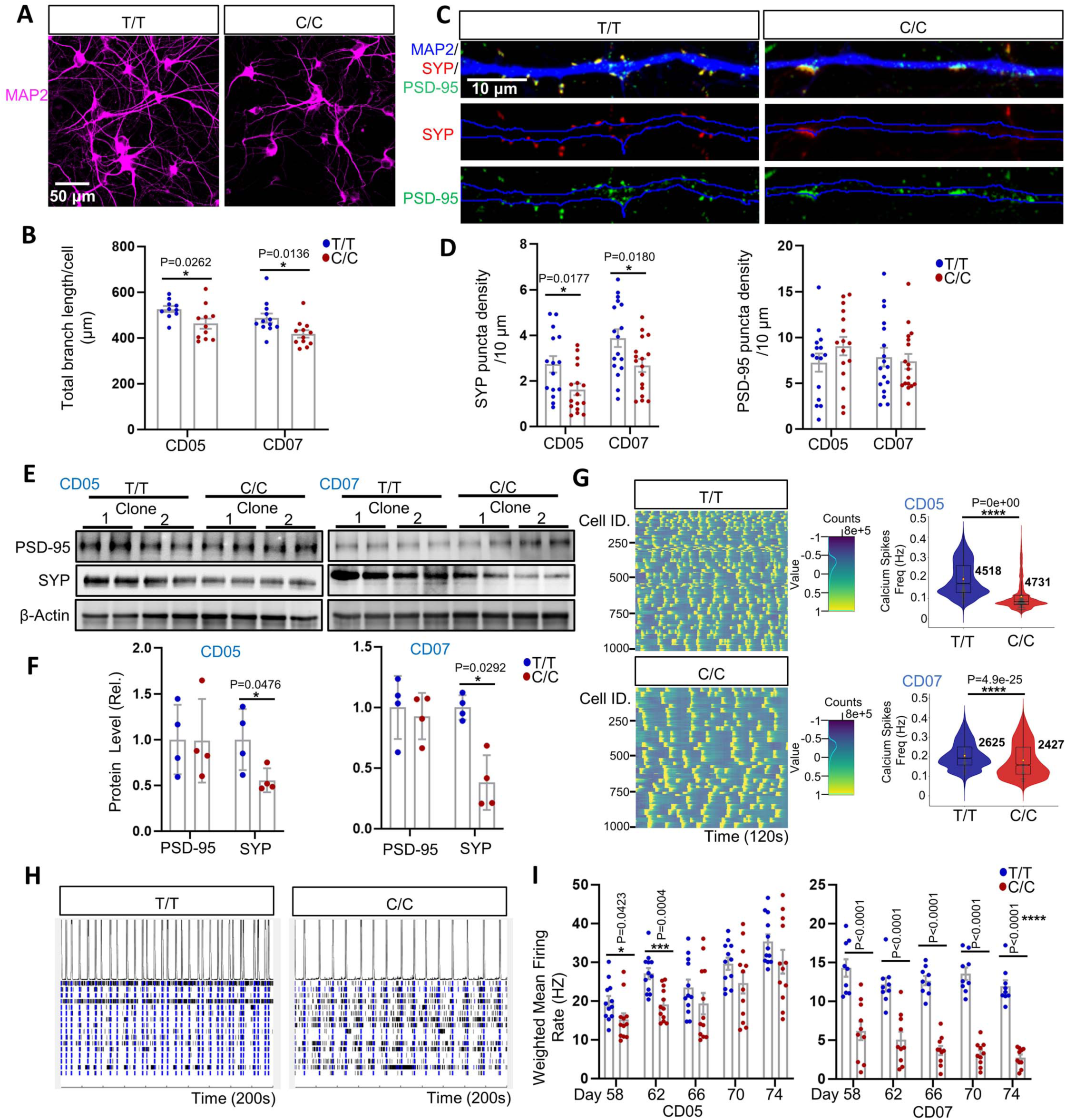
iGlut carrying the AD protective allele of rs1532278 at the *CLU* locus are more morphologically complex and functionally active. (A) MAP2 staining for analyzing iGlut dendritic branches in day-30 iGlut-mAst co-cultures. (B) Quantification of total branch length per cell (by Cellprofiler). n=10-12 coverslips per group from two independent experiments (5-6 coverslips per clone, 2 clones per line; 3-4 field of views, FOVs, averaged per coverslip). (C) MAP2, SYP, and PSD-95 staining for assaying synaptic puncta. (D) Quantification of SYP and PSD-95 puncta density per 10 µm. n=15-17 neurons per group (1-2 neuron per coverslips, 5-6 coverslips per clone, and 2 clones per line). (E-F) Western blotting for measuring PSD-95 and SYP levels in day-30 iGlut-mAst co-cultures. n=4 samples per group (2 clones per line and 2 biological replicates per clone). (G) Calcium imaging shows high firing frequency with T/T iGlut in co-cultures. Left panel, heatmaps of neuron firing peaks in calcium imaging assay during 120 seconds of recording (representative image from 1000 cells of CD05); Right panels, quantification of neuron firing frequency. The number of neurons assayed is shown in the violin plot (from 2-3 clones per line with 6 replicate wells per clone). (H) Representative roster blot of MEA from the CD07 line (a segment of 200 seconds is shown). (I) Weighted mean firing rate in MEA. n=9-12 wells per group (2-3 clones per line and 3-4 wells per clone). Violin plots are shown with box and whisker, all other statistical graphs depict mean±SEM. Scale bars are indicated in the corresponding images. * *p*<0.05, ** *p*< 0.01, *** *p*< 0.001, and **** *p*< 0.0001.

Next, we performed calcium imaging and multi-electrode array (MEA) analyses for iGlut/mAst co-cultures to examine the effect of the AD-protective *CLU* allele on neuronal electrophysiological properties. We observed a higher neuron calcium transmission (or firing frequency) for the T/T versus the C/C iGlut (**Fig. 2G**). Concordantly, we found that neurons with T/T alleles showed an elevated weighted mean calcium spike rate and more frequent network bursts in MEA (**Fig. 2I and S4D**). We also observed stronger neuronal network synchronicity in MEA for T/T alleles (**Fig. S4E**). These findings suggest that the AD protective allele of *CLU* is associated with more neuronal excitation. Given that neuronal hypoactivation is a notable characteristic of late-stage AD ^54^, our findings suggest that the AD protective allele of *CLU* may confer resilience to AD by promoting synaptic maturity and neuronal activity.

### Neuronal CLU mediates the effect of the AD protective allele of rs1532278 on neuronal excitability

To validate whether the enhanced neural excitability in iGlut carrying the AD protective allele of rs1532278 was due to the increased expression of *CLU*, we used CRISPR/Cas9 to introduce a ∼200 bp homologous deletion in the OCR that flanks rs1532278 without disrupting the flanking exons in both iPSC lines (**Fig. 3A and S5A**). We then differentiated these OCR-deleted (OCR-del) iPSCs into iGlut and cultured them with or without mAst. We found that the homozygous OCR deletion significantly reduced both *CLU* mRNA and sCLU protein levels compared to the non-edited T/C lines, regardless of the presence of mAst in the culture (**Fig. 3B, S5B and C**). Consistent with reduced *CLU* expression being detrimental to neuron physiology, SYP levels were reduced in the OCR-del iGlut (**Fig. 3C, 3D, S5E and F**). Neuronal excitability was also impaired in iGlut carrying the OCR-del, as evidenced by decreased neuron calcium spike (firing) frequency in the calcium imaging experiment (**Fig. 3E)** and reduced weighted mean firing rate (**Fig. 3F**), number of bursts, and synchronicity in the MEA experiment (**Fig. S5G and H**). These results support that the OCR flanking the ASoC SNP rs1532278 contains an enhancer sequence (**Fig. 1B**) that modulates *CLU* expression, thereby altering neuronal excitability.

**Figure 3.**
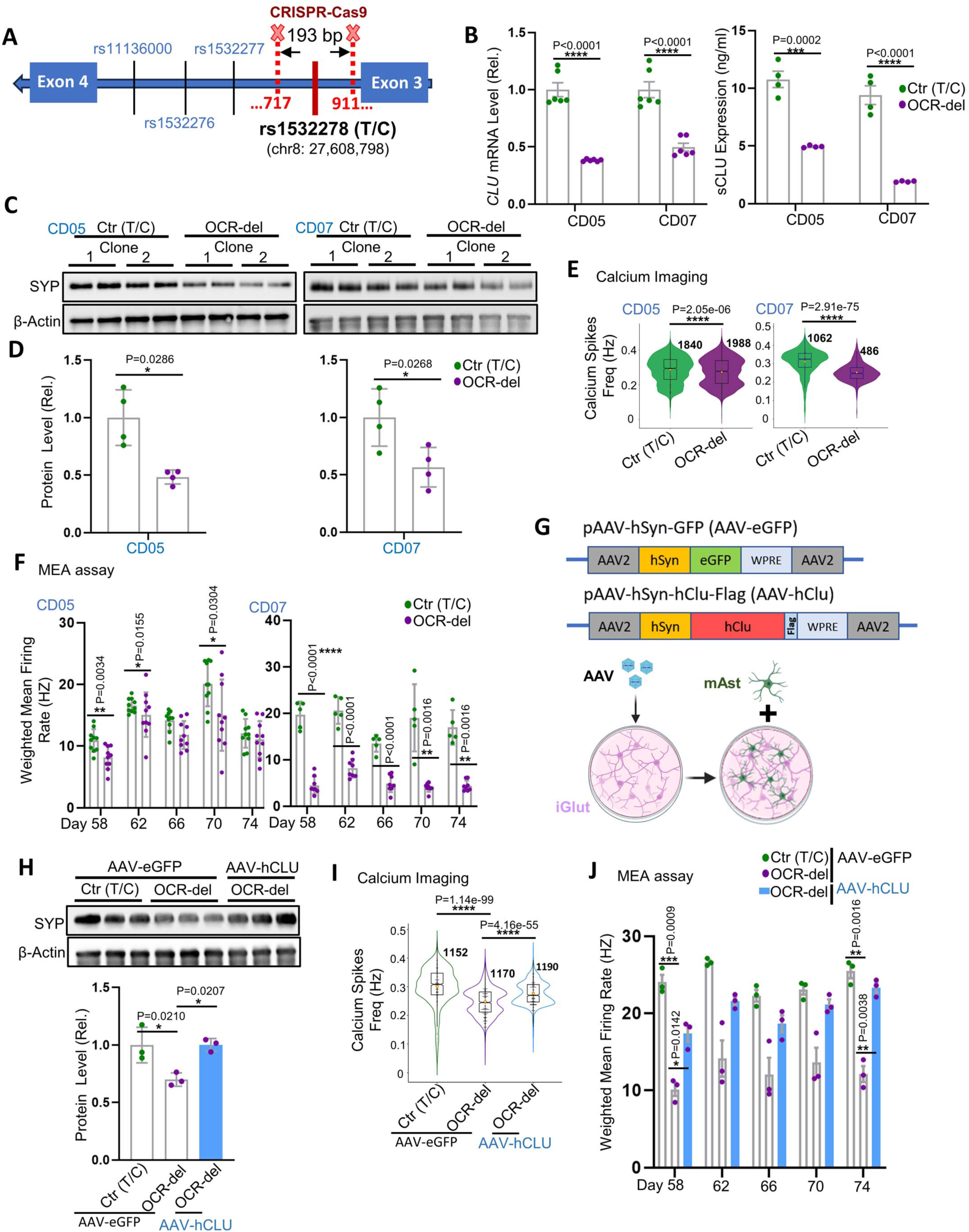
Neuronal CLU mediates the AD protective allele effect of rs1532278 on neuron excitability. (A) Schematic diagram of the rs1532278-flanking OCR deletion by CRISPR-Cas9. (B) *CLU* mRNA level (n=6 per group, 2 clones per line, and 3 replicates for each clone) and sCLU level (n=4 per group, 2 clones per line, and 2 replicates for each clone) in day-30 iGlut-mAst co-culture. (C-D) Western blotting detected SYP levels in day-30 iGlut-mAst co-cultures. n=4 per group. (E) Fire frequency analysis of calcium imaging assay. The indicated number of neurons is from 2 clones of each line and 3 replicated wells per clone. (F) Weighted mean firing rate analysis in MEA. n=5-10 wells per group (2-5 wells per clone and 2 clones per line). (G) The schematic diagram illustrates CLU overexpression in iGlut. AAV-hCLU, hCLU-Flag cDNA was inserted into the vector to replace the eGFP sequence in AAV-eGFP. AAV Virus infected neurons on day 9, followed by replating neurons with mAst on day 12. For the calcium imaging assay, the jRCaMP1b virus was introduced on day 10. (H) Western blotting for SYP in day-30 iGlut-mAst co-cultures. n=3 samples per group (3 biological replicates from one clone of the CD07 line). (I) Calcium transmission frequency. The number of neurons from 5 replicated wells in each group from one clone of line CD07. (J) Weighted mean firing rate in MEA. n=3 wells per group from one clone of line CD07. Violin plots are shown with box and whisker; all other graphs depict mean±SEM. Scale bars are indicated in the corresponding images. * p<0.05, ** p< 0.01, *** p< 0.001, and **** p< 0.0001.

To further confirm that neuronal excitability is modulated by CLU, we infected neurons with AAV-*CLU* (human) virus to rescue *CLU* expression in iGlut derived from iPSC line (CD07) homozygous for the rs1532278-flanking OCR-del. We examined SYP levels using WB and assayed neuronal excitability using calcium imaging and MEA experiments in AAV-infected neurons co-cultured with mAst (**Fig. 3G-J and S5I-K**). We found that restoration of *CLU* expression rescued SYP levels (**Fig. 3H**) and neuronal excitability in iGlut infected with AAV-*CLU*, as evidenced by the increased firing frequency in calcium imaging, higher weighted mean firing rate, more frequent network bursts, and better synchronicity compared to the control (AAV-eGFP) group (**Fig. 3I-J and S5K**). Hence, our data indicate that neuronal CLU mediates the effect of the AD protective allele of rs1532278 on neuronal excitability.

### Transcriptomic analysis supports an enhanced neuronal excitation and altered neuron-astrocytic lipid metabolism by the AD protective allele of rs1532278

To gain insights into the underlying molecular mechanisms that functionally link the AD protective T allele of rs1532278 with neuronal excitability, we conducted RNA-seq of iGlut/mAst co-cultures of the isogenic CRISPR-edited iPSC lines carrying the AD protective alleles T/T vs. risk alleles C/C. We bioinformatically separated the RNA-seq reads of iGlut (human origin) and mAst (mouse origin) using Salmon program **(Fig. 4A)** ^55^. To confirm the validity of the human iGlut RNA-seq reads, we evaluated the transcriptomic similarity of the iGlut to that from our previous scRNA-seq data of iGlut from the same iPSC lines ^42^ and the scRNA-seq data of different cell types of postmortem brain ^23^. We found a moderate to strong positive correlation between the expression of our co-cultured iGlut and of previous scRNA-seq datasets (r=0.49-0.64), supporting the validity of our bioinformatically separated RNA-seq reads of iGlut and mAst (**Fig. S6A and Table S3**). Furthermore, principal component analysis (PCA) of all the iGlut (n=40,162 transcripts) and mAst (n=32,476 transcripts) samples with different alleles of rs1532278 showed that RNA-seq samples were clearly grouped by rs1532278 genotypes (**Fig. S6B**), suggesting a pronounced transcriptional difference in iGlut with the AD protective allele T /T vs. C/C of rs1532278. Interestingly, mAst also exhibited transcriptomic differences based on their exposure to T/T iGlut or C/C iGlut, suggesting allele-specific neuron-glia interactions.

**Figure 4.**
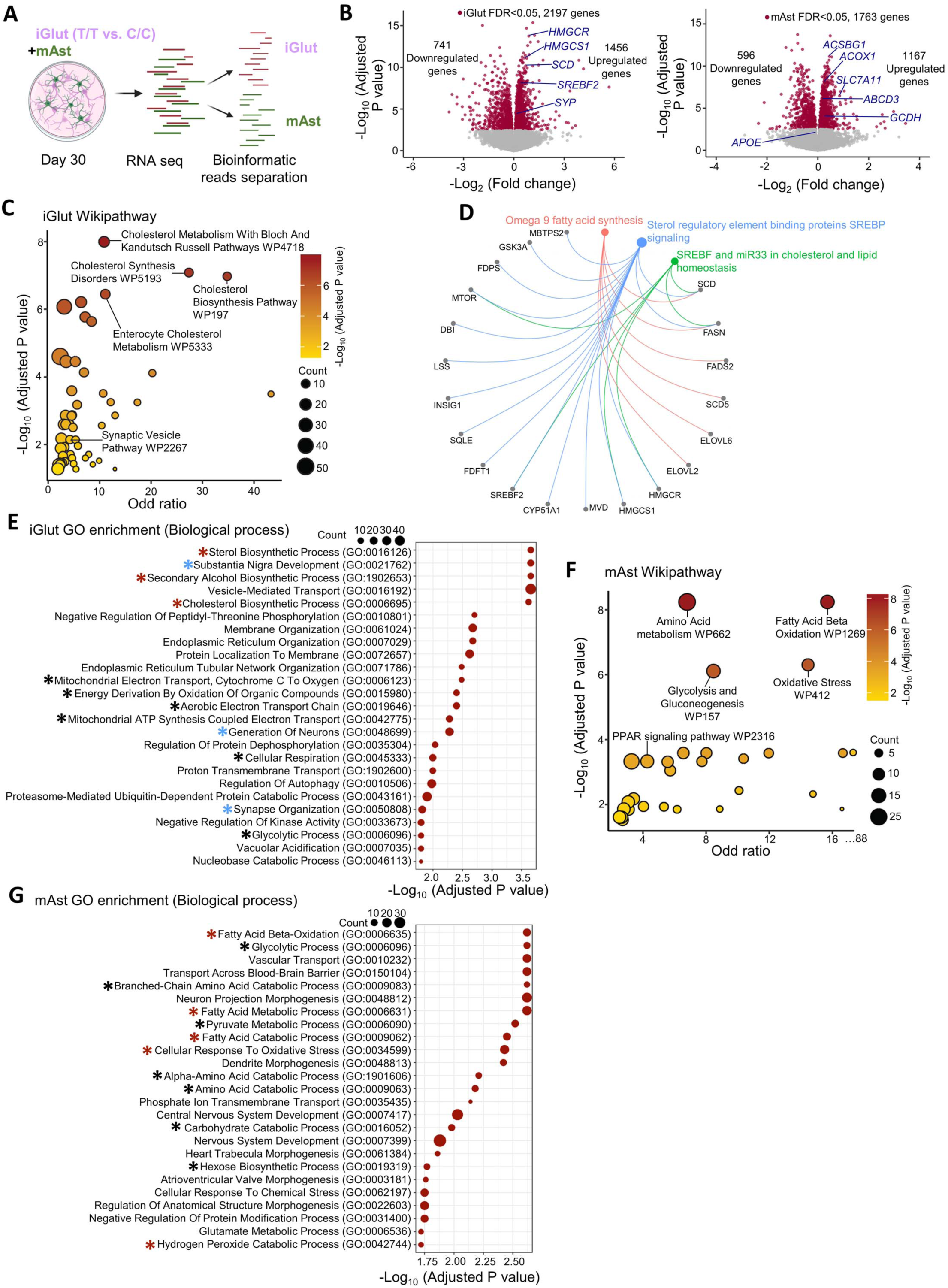
Transcriptomic analysis of iGlut-mAst co-cultures highlights the involvement of lipid metabolism. (A) Schematic diagram for RNA-seq data processing pipeline to separate sequencing reads of human and mouse origins. (B) Volcano plots show DE genes in iGlut (left) and the co-cultured mAst (right) with highlighted genes. Log_2_FC was based on a comparison of T/T vs. C/C cultures. (C) Wiki pathway analysis of upregulated gene sets in iGlut. (D) The Cnet plot depicts the enriched fatty-acid biosynthesis pathways and the upregulated genes. (E) GO-term (biological process) enrichment in significantly upregulated gene sets in iGlut. Red star, lipid biosynthesis pathways; black star, energy metabolism pathways; blue star, neuron-morphology related pathway. (F) Wiki pathway analysis of the significantly upregulated gene sets in mAst. (G) GO-term (biological process) enrichment in significantly upregulated gene sets in mAst. Red star, fatty acids metabolism-related pathways; black star, metabolism-related pathways.

We next analyzed for differential expression (DE) of genes in iGlut and mAst between the isogenic pairs of co-cultures (T/T vs C/C) (**Fig. 4B, Tables S4 and 5**). For iGlut carrying the AD protective alleles T/T, we identified 1,456 upregulated and 741 downregulated genes (FDR<0.05) (**Fig. 4B**). Wiki pathway analysis ^56^ of the top-ranking DE genes (P<0.001) further showed that 49 gene pathways were significantly enriched in the upregulated gene sets (**Fig. 4C, Table S4**), and 14 in the downregulated gene sets (adjusted P value<0.05) (**Table S5**).

The synaptic vesicle pathway was among those upregulated, including the presynaptic gene *SYP* whose expression in T/T iGlut was relatively higher than that of C/C as evidenced by immunofluorescence staining and WB analysis (**Fig. 2C-F**). In addition, we also found synapse, axon and neuron projection, and development related pathways enriched in both up- and downregulated genes **(Fig. 4E and S6D**, **Tables S10 and S11)**. Unexpectedly, we found that the upregulated genes were enriched for AD related pathways including APP secretase genes *PSEN1* and *BACE1* (**Table S6**), which prompted us to measure the secreted Aβ levels, including Aβ (1-40) and Aβ (1-42), in our iGlut/mAst co-cultures by ELISA. We found increased levels of both Aβ peptides in the culture media of T/T iGlut/mAst compared C/C iGlut/mAst co-cultures (**Fig. S6C**), which may be a result of an enhanced neuronal activity as previously reported ^57,58^. These results provide mechanistic insight into how the AD protective allele T of *CLU* increases neuronal excitation (**Fig. 2G-I**).

Interestingly, we found that the most enriched gene pathways are related to fatty acid (FA) and cholesterol biosynthesis (**Fig. 4C** and **D, Table S6**). The notable upregulated genes in T/T neurons included *HMGCR* (*3-Hydroxy-3-Methylglutaryl-CoA Reductase*; 3^rd^ most upregulated; log_2_FC=0.89, FDR= 1.20 × 1010^−10^) that encodes a rate-limiting enzyme for cholesterol synthesis, and *SCD* (24^th^ most upregulated; log_2_FC=0.70, FDR= 8.13 × 1010^−8^) that encodes *Stearoyl-CoA Desaturase* that is involved in FA biosynthesis. Moreover, energy metabolism pathways such as metabolic reprogramming, glycolysis, gluconeogenesis, and oxidative phosphorylation were among the top-ranked enriched Wiki pathways (**Table S6**). Gene Oncology (GO) analysis showed consistent results with Wiki pathway analysis: the top 25 enriched GO terms (biological processes) are strongly enriched for genes involved in lipid/cholesterol synthesis (**Fig. 4E and Table S10**).

Although enhanced neuronal excitability tends to require more energy consumption ^59^, neurons cannot sufficiently catabolize FAs as an energy source. Hence, neurons typically transfer FAs to surrounding astrocytes for catabolism, providing energy to neurons ^60–62^. We thus examined whether the elevated neuronal CLU in T/T iGlut might have transcriptional effects on lipid metabolism in the co-cultured mAst. Our DE analysis identified a total of 1,167 upregulated and 596 downregulated genes in mAst (FDR<0.05) **(Fig. 4B)**. Wiki pathway analysis of the top-ranking DE genes (*p*<0.001) further identified 31 significant pathways enriched in the upregulated gene sets (**Fig. 4F, Table S8**), and 29 in the downregulated gene sets in mAst (adjusted p value<0.05) (**Table S9**). Interestingly, the most enriched Wiki gene pathways, which were also supported by GO enrichment analysis, were those related to energy metabolism (**Fig. 4F and G, Tables S8 and S12**), e.g., FAs β-oxidation/degradation, oxidative stress, peroxisome proliferator-activated receptor (PPAR) signaling pathways, and glycolysis (**Fig. 4F, Tables S8 and S12**). FAs β-oxidation is recognized as a primary source of ROS and is commonly linked to oxidative stress ^63^, and a homeostatic balance of cellular ROS level has been considered essential for maintaining normal cellular physiological function ^64^. It is noteworthy that PPAR signaling pathways play a crucial role in systemic cell metabolism and energy homeostasis ^65^, and one of the proteins in this pathway, ACSBG1 (Acyl-CoA Synthetase Bubblegum Family Member 1), enables long-chain FA-CoA ligase activity and very long-chain FA (VLCFA)-CoA ligase activity (MGI:2385656) and *ACSBG1* was the second-most upregulated (log_2_FC=0.46, FDR=3.39 × 1010^−8^) gene in mAst co-cultured with T/T iGlut **(Fig. 4B)**. Peroxisomes are indeed a source of ROS in glia and provide energy to neurons through β-oxidation of VLCFA ^66,67^. Independent from, but positively coupled with FAs β-oxidation in astrocytes, glycolysis also provides energy to neurons ^68–70^. It is thus conceivable that these processes in mAst are protective by providing more energy reserve for neuronal function. Taken together, our transcriptomic analyses of the iGlut/mAst co-cultures provide not only mechanistic support for the effects of the AD protective allele of rs1532278 at the *CLU* locus on enhanced neuron excitability, but also unravel potential roles of neuron-glia lipid transfer and metabolism in mediating the effects of this allele.

### Neuronal CLU augments neuron-glia lipid transfer and reduces neural LD accumulation

Following the interesting lead of the transcriptional effects of the AD protective allele on gene pathways related to lipid synthesis and metabolism in iGlut/mAst co-cultures, we hypothesized that the T/T iGlut and its associated increases in neuronal CLU production may promote LD formation in neurons and/or mAst to serve as energy storage ^71,72^. We first stained T/T or C/C iGlut for LDs using the neutral lipid dye LipidTox in the absence of mAst. Despite the upregulated lipids and cholesterol synthesis genes in T/T iGlut (**Fig. 4C-E**), we surprisingly observed that iGlut with this AD protective allele and higher *CLU* expression had fewer LDs compared with AD risk alleles C/C **(Fig. 5A and B, and S7A)**. To further validate this observation, we overexpressed *CLU* using AAV in iGlut (T/C at rs1532278). We found that similar to T/T iGlut which have high expression of *CLU* compared to C/C iGlut, the AAV-*CLU* group also showed reduced LDs **(Fig. 5C and D, and S7B)**.

**Figure 5.**
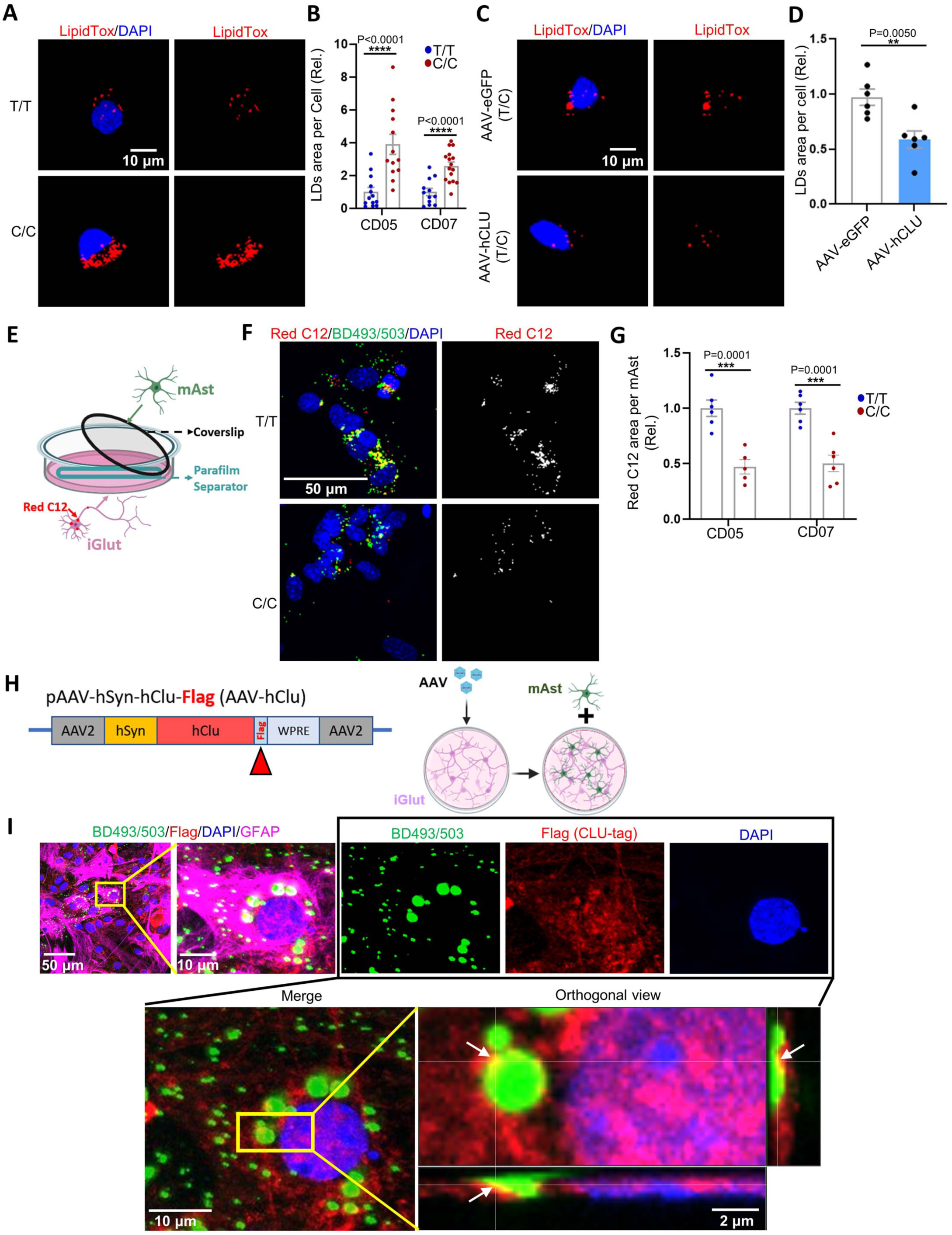
Neuronal CLU facilitates neuron-to-astrocyte lipid transfer. (A-B) Lipid droplet (LD) staining by LipidTox in day 25 iGlut pure cultures (T/T vs. C/C). n=12 coverslips per group from three independent experiments (2 clones per line, 6 coverslips for each clone with 3-5 cells averaged from each coverslip). (C-D) LipidTox staining of Day 25 iGlut pure cultures after AAV-CLU overexpression. n=6 coverslips per group (one clone of unedited CD07 with genotype T/C, 3-4 cells averaged from each coverslip). (E) The schematic diagram for the Red C12 lipid transfer assay. (F-G) Red C12 and LDs (BD493/503) signals in mAst. n=5-6 coverslips per group (mAst co-cultured with 2 clones per line, 2-3 coverslips for each clone with 3-5 FOV averaged from each coverslip). (H) The schematic diagram for overexpression of CLU tagged with Flag. (I) Co-localization of neuron secreted and AAV-derived CLU (Flag+) and LDs (BD493/503+) in mAst from day-30 iGlut-mAst cocultures after AAV-hCLU infection (same experimental approach as Fig. 3G). Data, mean±SEM. * *p*<0.05, ** *p*< 0.01, *** *p*< 0.001, and **** *p*< 0.0001.

*CLU* encodes an apolipoprotein, APOJ, which may have a similar lipid transferring functions as APOE ^62,73^. Previous studies have shown APOE function to directly impact the transfer of lipids from neurons to astrocytes where the lipids are incorporated into glial LDs ^10,61,62^. Hence, we reasoned that more *CLU* expression from T/T iGlut may promote this process of lipid transfer from neurons to glia. We conducted a lipid transfer assay ^10,74^ by first pre-labeling iGlut with a fluorescently labeled FA probe (Red C12) and then co-culturing these Red C12-loaded iGlut with mAst in the same well without direct physical neural-glia contact (**Fig. 5E**). This system allows the neuronally sourced Red C12 to be transferred to mAst upon the induction of stress where they are incorporated into LDs ^10,62,75^. We observed a 2-fold increase of the Red C12 positive LDs in mAst co-cultured with T/T iGlut compared to mAst co-cultured with C/C iGlut (**Fig. 5F and G**). Culturing with T/T iGlut did not affect *Apoe* expression in mAst (**Fig. 4B**), suggesting that the increase of the Red C12 in mAst is attributed to the elevated neuronal CLU expression mediated by the AD protective allele. These data may explain the lack of neuronal LD in the T/T iGlut (**Fig. 5A-B**) as more lipids may have been expelled from these iGlut due to high CLU expression, leading to very few neuronal LDs but many astrocytic LDs. Notably, neurons have a limited ability to catabolize FAs, and the LD accumulation in neurons is usually considered detrimental to neuronal function ^60,62^. These data indicate a CLU-mediated neural protective mechanism of the T/T allele of the AD associated SNP rs1532278.

To corroborate that neuronal CLU can associate with lipid particles and carry lipids from neurons to astrocytes, we first infected iGlut with AAV to overexpress Flag-tagged CLU and co-cultured these AAV-infected iGlut with mAst. We then stained for LDs with the neutral lipid dye BODIPY (BD493/503) in mAst (GFAP+) to examine the co-localization of LDs with neuronally sourced CLU. We found that the Flag-tagged neuronal CLU was able to enter mAst and co-localize with BD493/503 positive LD (**Fig. 5H**). These results support a previously unappreciated role of neural CLU in neuron-to-glia lipid transfer. Further, these data are consistent with the idea that reducing neural LD accumulation is neuroprotective and that higher CLU expression associated the protective rs1532278 allele is promoting neuron-glia interactions in the context of LD accumulation.

### Neuronal CLU promotes LD formation, energy preservation, and ROS production in mAst

We next examined whether total LD amount was altered in mAst as a result of the increased neuronal CLU when co-cultured with iGlut carrying the AD protective T/T allele (**Fig. 6A and S7C**). We first quantified the relative LD distributions in iGlut versus mAst in the co-cultures. We initially stained the co-cultured cells with LipidTox and identified iGlut versus mAst using the neuron-specific marker MAP2 (**Fig. 6A and S7C**). We found that while iGlut occupied ∼40% of the immunostaining region (MAP2 volume / [MAP2+GFAP volumes]), neural LDs only accounted for ∼5% of the total LDs (**Fig. 6A**). The vast majority of LDs were found in mAst (∼95%) (**Fig. 6A**), consistent with neural-to-glia lipid transfer being augmented by neuronal CLU (**Fig. 5E-G**) and with previous reports in which LDs *in vivo*, or in the presence of glial cells, are predominantly present in glia ^76–78^. Next, we compared the allelic effect of the AD risk SNP rs1532278 on LDs in iGlut/mAst co-cultures. We found that T/T iGlut caused a significant increase in total LDs, predominantly within the mAst, compared to C/C iGlut (**Fig. 6B and C, and S7D**). CLU overexpression by AAV in iGlut, followed by co-culture with mAst, also showed an elevation of LDs within mAst (**Fig. S7E and F**), demonstrating that the increase in total astrocytic LDs was due to a higher expression of CLU.

**Figure 6.**
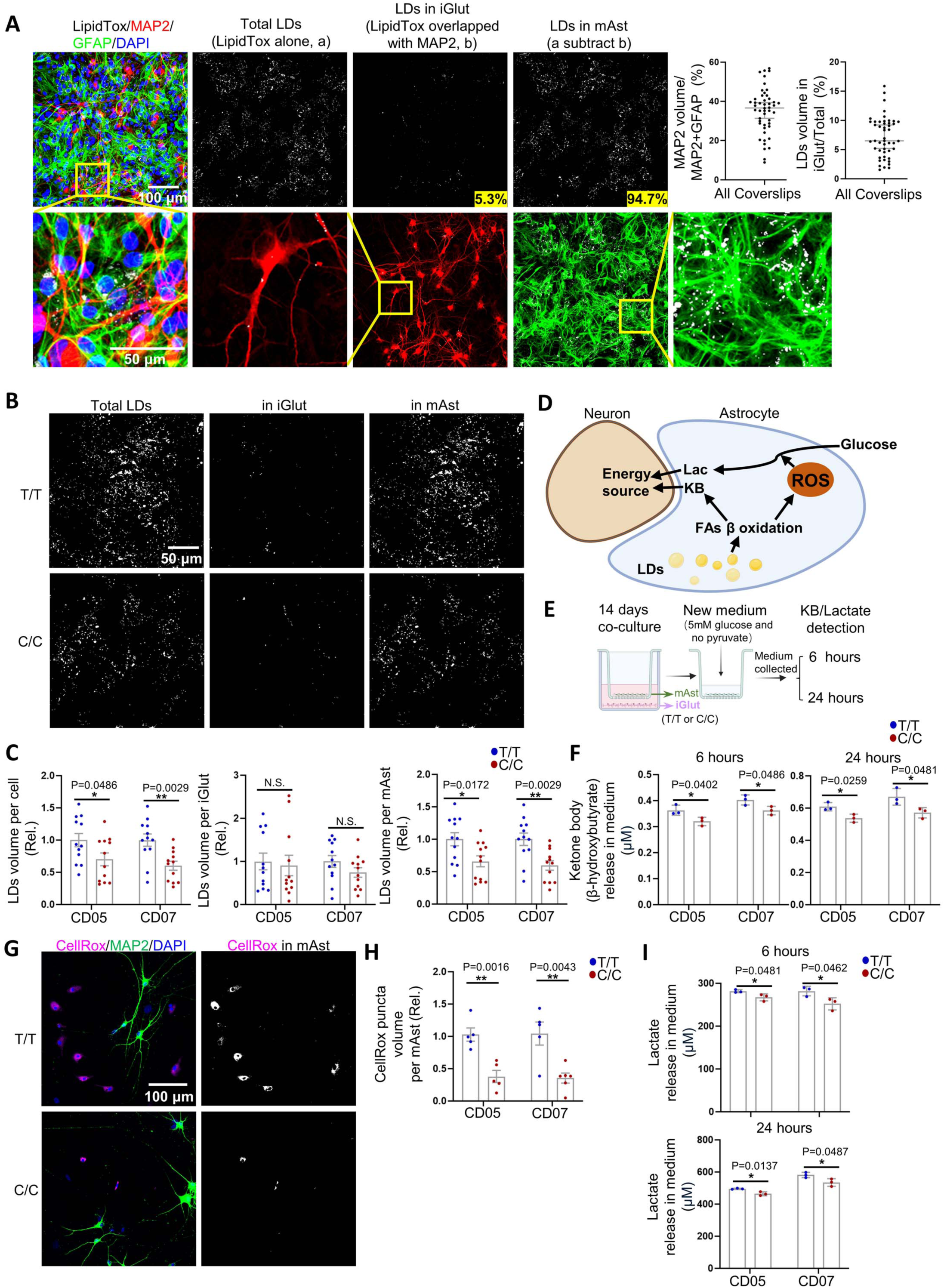
The AD protective allele of rs1532278 at the *CLU* locus in iGlut induces LD accumulation and produces more ROS in co-cultured mAst. (A) LD staining in iGlut-mAst co-cultures by LipidTox (Example images of C/C CD07 co-cultures). LDs were categorized by neuronal (MAP2+) or astrocytic (i.e., not neuronal) and quantification was shown in the right panels. Neuronal LDs, LDs in iGlut relative to total LD volume. The proportion of the image area occupied by iGlut was calculated as MAP2 volume divided by the total of GFAP and MAP2 volume. n=48 coverslips from all genotypes of both CD05 and CD07 lines (two independent experiments). (B-C) LD staining and quantification by iGlut genotype in different cell types in day-30 iGlut-mAst co-cultures. n=12 coverslips per group from two independent experiments (2 clones per line, 6 coverslips for each clone with 2-4 FOV averaged from each coverslip, shown are example images of CD07). (D) The schematic diagram depicts the FA β oxidation process of astrocytes, where they produce KB (ketone body) and ROS; the glycolysis is further facilitated by ROS and generates Lac (lactate); both KB and Lac are further transferred to surrounding neurons as the energy source. (E) the schematic diagram depicts the transwell co-culture system to measure KB and Lac release from mAst. (F) The amount of ketone body (β-hydroxybutyrate) released from mAst at different time points in the transwell co-culture system. n=3 wells per group. (G-H) CellROX staining in iGlut-mAst co-cultures. n=5-6 coverslips per group from two independent experiments (2 clones per line, 2-3 coverslips for each clone with 2-4 FOV averaged from each coverslip, shown are example images of CD07). (I) The amount of lactate released from mAst at different time points in the transwell co-culture system. n=3 wells per group. Data, mean±SEM. * *p*<0.05, ** *p*< 0.01, *** *p*< 0.001, and **** *p*< 0.0001.

As a lipid storage depot, LDs can protect the cell from lipotoxicity and provide energy support ^79,80^. In the brain, FAs are initially synthesized in neurons and then transported to astrocytes where FAs are stored as LDs and later metabolized to replenish the brain’s energy supply playing a physiological role ^62,69–72^. As we observed increased LD formation in mAst co-cultured with iGlut carrying the protective T/T allele or overexpressing CLU (**Fig. 6B and C, and S7E and F**) and our transcriptomic analyses identified enriched gene pathways related to FAs β-oxidation in the T/T iGlut (**Fig. 4F and G**), we next examined whether the elevated LD level in mAst co-cultured with T/T iGlut was accompanied with increased FAs β-oxidation. FAs β-oxidation is known to produce ketone bodies (KB) as a neuronal energy source while generating ROS ^81^ (**Fig. 6D**). We next employed a commonly used transwell system ^10,69,70^ to examine KB specifically released from mAst, where we co-cultured iGlut carrying T/T or C/C alleles with mAst for 14 days. Subsequently, we took out the mAst to detect KB levels (β-hydroxybutyrate) after 6 and 24 hours in fresh medium (**Fig. 6E**). We found mAst co-cultured with T/T iGlut exhibited higher KB levels at both time points, suggesting more FAs were catabolized to ketone to likely support higher energy demands of T/T iGlut (**Fig. 6F**). Moreover, because lactate is a well-established neuronal energy substrate derived from glycolysis that is coupled with FA β-oxidation in astrocytes ^67,69,70^ and because it was transcriptionally enriched in mAst co-cultured with T/T iGlut (**Fig. 4F and G**), we also assayed the lactate release from the mAst in the same transwell system (**Fig. 6D**) and observed an increase in lactate levels in mAst co-cultured with T/T iGlut (**Fig. 6I**). Finally, as FA β-oxidation also generates ROS (**Fig. 6D**), we employed CellROX staining to directly visualize ROS levels within iGlut/mAst co-cultures and used neuron-specific MAP2 staining to distinguish neuronal from astrocytic CellROX signals. We observed that almost all CellROX signals were detected in mAst but not in iGlut (**Fig. 6G and H, and S7G**), indicating a much lower neuronal ROS level when co-cultured with mAst. Secondly, we found that mAst co-cultured with T/T iGlut gave rise to significantly higher CellROX signals than when cultured with isogenic C/C iGlut (**Fig. 6G and H, and S7G**). Taken together, these results suggest the increased expression of neural CLU caused by the T allele along with the higher neuronal activity of T/T iGlut leads to elevated levels of astrocytic LDs, likely serving as energy storage and protecting against neuronal oxidative damage ^61,75^.

### LD-accumulated astrocytes show reduced glutamate uptake

Although the AD protective allele of rs1532278 at the *CLU* locus may promote neuronal excitability independently from facilitating the astrocytic LD formation, it is also plausible that these two processes are intrinsically coupled. In fact, astrocytes are also known to play an important role in maintaining glutamate homeostasis and coordinating neuronal excitation in the brain ^82^, while ROS have been shown to reduce glutamate uptake in AD fibroblasts ^83^. We thus hypothesized that the neuronal CLU-induced LD accumulation and ROS production in the co-cultured mAst may reduce glutamate uptake by astrocytes and thus promote neuron excitability. To test this hypothesis, we employed a commonly used transwell system ^10^, where we co-cultured iGlut carrying T/T or C/C alleles with mAst for 14 days. Subsequently, we removed the inserts with mAst to conduct a glutamate uptake assay (**Fig. 7A**). We found that T/T iGlut/mAst co-cultures had a ∼20% higher amount of residual glutamate in the culture media compared to C/C iGlut/mAst co-cultures (**Fig. 7A**). To determine whether the reduction of glutamate uptake of mAst was attributed to higher ROS level in the mAst co-cultured with T/T iGlut, we applied an antioxidant, NACA (N-acetylcysteine amide), to reduce ROS in the iGlut/mAst transwell co-cultures and assayed the glutamate uptake. We found that ROS inhibition significantly enhanced glutamate uptake of mAst co-cultured with T/T iGlut and to a lesser extent for mAst co-cultured with C/C iGlut (**Fig. 7B**). These results suggest that the glutamate uptake function of mAst is reduced through LD-enhanced ROS production when co-cultured with iGlut carrying the AD protective allele, possibly to sustain neuron excitability.

**Figure 7.**
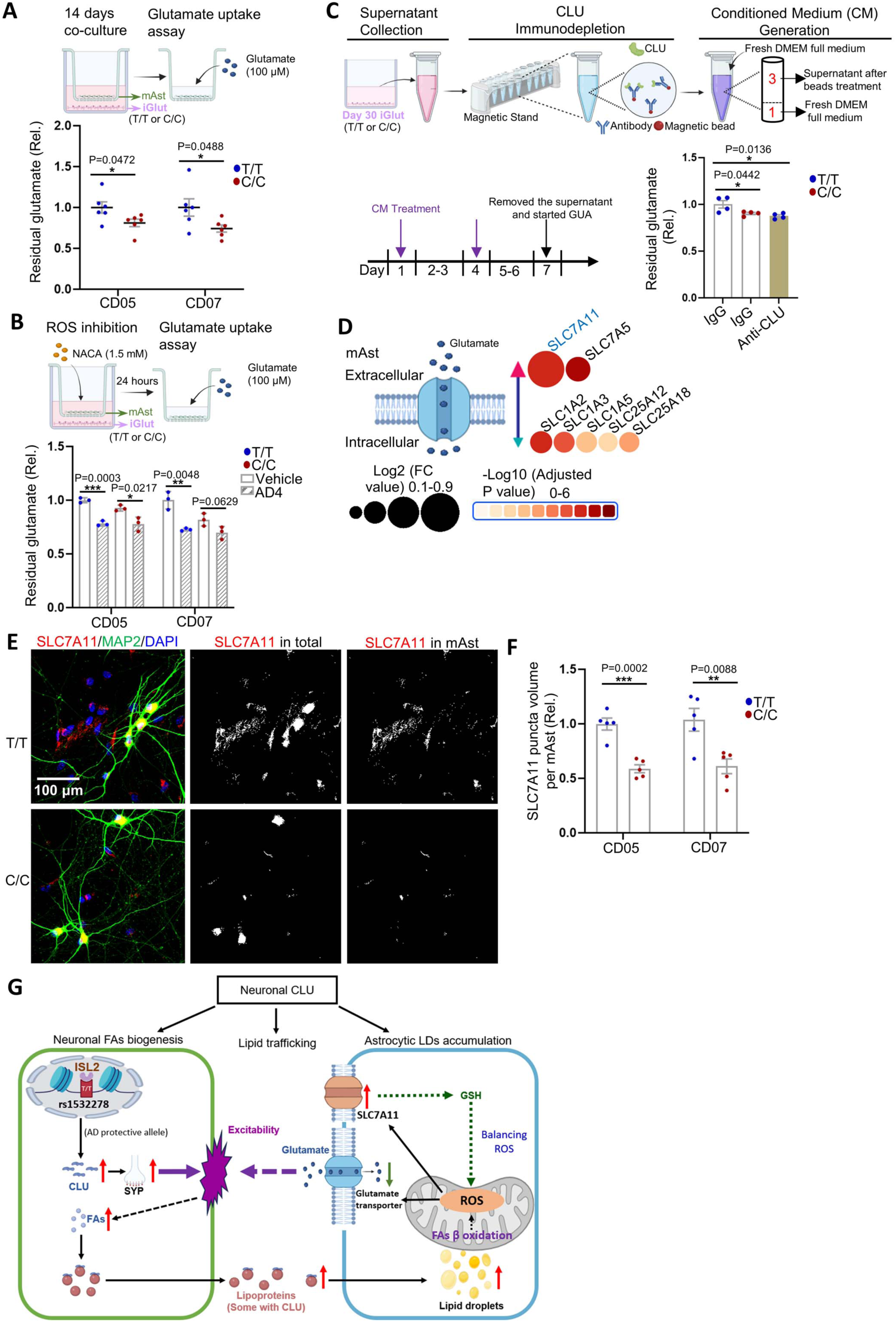
Astrocytes co-cultured with iGlut carrying the AD protective allele show reduced glutamate uptake. (A) Neural allelic effect on mAst glutamate uptake diagram showing the transwell coculture system for glutamate uptake assay in mAst (upper panel) and the quantification (down panel). n=6 wells per group (2 clones per line, 3 wells for each clone). (B) The schematic diagram (upper panel) shows the test for whether ROS inhibition can affect glutamate uptake in mAst in the transwell co-culture system; quantification of glutamate uptake assay after ROS inhibition in mAst. n=3 wells per group. (C) CLU immunodepletion assay confirms the role of CLU in glutamate uptake by mAst. Up panel, the diagram of the CLU immunodepletion assay; bottom-left panel, the culturing and assay timeline; bottom-right panel, the quantification of residual glutamate in glutamate uptake assay. n=4 wells per group. Note the similar levels of residual glutamate of the C/C group with CLU-depletion (C/C; IgG) and the T/T group upon CLU-depletion (T/T; Anti-CLU). (D) All DE genes related to glutamate transfer in mAst. The arrow indicates the direction of glutamate transfer. (E-F) SLC7A11 staining of iGlut-mAst co-cultures (images of CD07 line). n=5 coverslips per group (2 clones per line, 2-3 coverslips for each clone with 3-5 FOV averaged from each coverslip). (G) A graphic model of how the AD protective allele promotes neural CLU expression, subsequently facilitating neuron-glia lipid transfer and fine-tuning mAst glutamate uptake, thereby maintaining neuronal excitability. Data, mean±SEM. * *p*<0.05, ** *p*< 0.01, *** *p*< 0.001, and **** *p*< 0.0001.

To further determine the role of CLU in mediating glutamate uptake, we carried out a CLU-immunodepletion assay. We collected supernatant from day-30 iGlut and immunodepleted CLU protein/lipoparticles from the supernatant of T/T or C/C iGlut (**Fig. 7C**). Subsequently, we reconstituted conditioned medium (CM) by adding fresh DMEM full medium, which is needed for maintaining mAst, into the immunodepleted supernatant. We then measured glutamate levels in mAst cultured with this CLU-depleted CM for seven days versus mAst grown in non-CLU depleted CM (**Fig. 7C**). We observed that in normal CM, mAst treated with CM of T/T iGlut exhibited higher residual glutamate compared to those treated with C/C iGlut CM (i.e., T/T showed less glutamate uptake). However, with CLU depletion (anti-Clu), the glutamate uptake function of mAst co-cultured with T/T iGlut was at a similar level compared to the mAst co-cultured with C/C iGlut (**Fig. 7C**). These results indicate that sCLU produced from iGlut, which facilitates neuron-to-glia lipid transfer, is a key player in mediating the protective allelic effect of CLU on the glutamate uptake function by mAst.

We further explored possible mechanisms that may underlie the reduced glutamate uptake function of mAst co-cultured with iGlut carrying the AD protective allele. We examined the transcriptomic changes for DE genes related to glutamate metabolism (**Table S5**) and found the upregulated genes in mAst co-cultured with T/T iGlut were highly enriched for the glutathione (GSH) metabolism pathway (**Fig. S8A and Table S8**). GSH serves as an intracellular antioxidant, neutralizing ROS and acting as a protective factor to reduce oxidative stress ^84^. GSH can also be shuttled from astrocytes to neurons, protecting them from oxidative damage ^70^. Hence, the upregulation of GSH metabolism may represent a compensatory mechanism in mAst to balance excess ROS production and maintain cell health. Interestingly, *SLC7A11*, a gene that encodes a cystine/glutamate antiporter that acts upstream of the GSH metabolism pathway ^85^, was the most transcriptionally upregulated gene among all glutamate transporters (**Fig. 7D**). We also independently confirmed its upregulation in mAst co-cultured with the AD protective allele (**Fig. 7E and F**). Taken together, these results suggest that elevated level of LDs coupled with ROS production in astrocytes co-cultured with T/T iGlut may promote neuronal excitability by fine tuning astrocytic glutamate uptake and/or export **(Fig. 7G)**.

## Discussion

Both *APOE* and *CLU* encode apolipoproteins and are associated with AD risk. However, compared to *APOE*, the functional connection of *CLU* with AD pathogenesis has been heretofore obscure. The challenge is determining the disease causal variants among multiple AD risk SNPs at the *CLU* locus that are in the noncoding portion of the genome as opposed to the *APOE4* risk allele which is a protein-coding SNP. Here, leveraging a systematic mapping for functional noncoding AD risk variants that alter chromatin accessibility (i.e., showing ASoC) in iPSC-derived neurons and glial cells ^46^ (**Table S1**), we identified rs1582273 (T/C) as a neuron-specific functional AD-associated variant. Further CRISPR/Cas9 editing of rs1582273 in iPSC-derived neurons showed that the AD protective allele T of rs1582273 specifically increased *CLU* expression in excitatory neurons (iGlut). We also found that elevated neuronal CLU helps promote neuronal excitability, neuron-to-glia lipid transfer, and lipid storage in the form of LDs within astrocytes. The accumulation of astrocytic LDs was associated with increased astrocytic ROS that reduced glutamate uptake, likely contributing to maintaining neuronal excitability. Our iPSC-based cellular modeling provides a mechanistic link between AD risk variants of *CLU* with CLU-mediated protective effects on neuron-glia communication to maintain homeostasis of lipid metabolism, energy consumption, and neuronal excitability.

Previous studies have primarily focused on astrocytic CLU due to its higher expression in astrocytes than in neurons ^29,86,87^. We found that the AD-associated rs1582273 of *CLU* is a neuron-specific functional SNP (ASoC), with its AD protective allele T *cis*-upregulating *CLU* expression specifically in iGlut **(Fig. 1**). This is consistent with another reported AD protective allele T of rs11136000 which also increases *CLU* expression. However, the association index for SNP rs11136000 suggests that it is not the functional SNP due to its strong linkage disequilibrium with other AD risk SNPs including rs1582273 (r^2^=0.98) that we identified as having functional consequences ^31–34^ (**Table. S1**). A recent study suggested that rs1582273 may function in astrocytes ^88^, whereas we show that rs1582273 specifically functions in neurons (iGlut) using SNP editing by CRISPR-Cas9 and ASoC mapping by ATAC-seq (**Fig. 1 and S3B**). While we do not exclude the possibility of rs1582273 having functional consequences in other cell types vulnerable to AD risk *in vivo,* our study strongly suggests that rs1582273 impacts neurons by altering expression at the *CLU* locus.

Plasma CLU has been shown to be protective by reducing neuroinflammation in an AD mouse model ^28^. Here, we report that the elevated neuronal *CLU* expression by the T allele of the AD-associated variant rs1582273 is also neuroprotective. The T allele is a derived allele and is a less frequent allele (∼38%) in the general population compared to the C allele. AD GWAS show that the T allele is less frequent in AD patients (odds ratio = 0.91), and, hence, we conclude that it is a protective allele. Although one could also interpret the C allele as detrimental given that its odds ratio in AD patients is 1.1, the C allele is less likely to confer significant AD risk in the general population, given its high population frequency. It is also interesting that the T allele of rs1582273 is only present in humans but not in other mammals (**Fig. S1A**), suggesting a potential evolutionary benefit of the T allele. Our observation of higher *CLU* expression in the cortex of controls vs. AD cases (**Fig. 1H**) and the results of our functional investigations also support a protective role of neuronal CLU. The main protective effects of neuronal CLU observed in our study include promoting neural excitability **(Fig. 2G-I**) and promoting lipid storage in the form of astrocytic LDs (**Fig. 6A-C and S7D**).

How does the elevated neuronal CLU promote neuronal excitation that is beneficial? CLU secreted from astrocytes has been reported to promote excitatory synaptic transmission in mice ^29^. CLU levels are positively correlated to synaptophysin expression ^89^ and neurite growth ^90^ in different models. In iGlut, we also observed that elevated CLU levels led to more elaborate dendrites and increased presynaptic sites (**Fig. 2A and B**). Such enhancement of neural structural maturity by CLU may promote neuron firing and synchronization. Although shorter neurites may exhibit lower cell capacitance and faster kinetics for neurons firing in certain AD models ^91,92^, longer neurites with enhanced synaptic density can feasibly amplify synaptic input signals and facilitate neuron firing. Importantly, we found that the enhanced neuronal activity by CLU correlated with enhanced neural CLU-mediated lipid transfer and LD metabolism when the neurons were co-cultured with astrocytes (**Fig. 5 and 6**). Furthermore, LD accumulation in mAst is accompanied by increased ROS accumulation that inhibits glutamate uptake (see below). As for the protective role of such CLU-mediated neuron excitability, we note that despite it being seemingly inconsistent with the commonly perceived clinical feature of neuron hyperexcitability in patients at early stages of AD, neuron hypoexcitability at a later stage is a common characteristic of neurodegeneration ^93,94^. In this regard, maintaining an appropriate level of neuronal excitability is pivotal for brain health, and hence potentially beneficial at a later stages of AD.

Here, we show that neuronal CLU promotes neuron-to-astrocyte lipid transfer and storage of neuronal lipids in the form of glial LDs. With an elevated CLU by the AD protective allele, neuronal FAs are transferred to surrounding mAst, resulting in LD storage in mAst (**Fig. 6A-C, and S7D**). The elevated LDs in mAst co-cultured with T/T iGlut could stem from either increased FA biosynthesis in iGlut coupled with the augmented neuronal CLU-mediated neuron-to-glia lipid transfer (**Fig. 4C-E and 5E-G**) and/or from reduced LD degradation in mAst. To this end, we noted that our transcriptomic data revealed that the lysosome was the top-ranking downregulated GO term (Cellular Component) in mAst **(Fig. S8B and Table S14**), suggesting that reduced lysosomal function may help maintain the level of LDs in a beneficial range. Interestingly, in contrast to the downregulation of lysosomal genes in mAst, genes related to lysosome and autophagy function in iGlut carrying the AD protective allele were upregulated (**Fig. S8B, Table S12 and S13**). Since the lysosome and autophagy pathways are crucial for regulating lipophagy, which breaks down LDs and maintains LD balance ^60^, an enhanced LD catabolism may also contribute to FA production in iGlut carrying the AD protective allele, which may synergistically contribute to LD formation in mAst.

Astrocytes catabolize FAs from LDs through β-oxidation to release energy, a process coupled with production of ROS ^60,61^. In support of the beneficial effect of the AD protective allele T on preserving LDs as an energy source, we found that more FAs were catabolized to ketones and there was an elevated lactate release in mAst co-cultured with T/T iGlut (**Fig. 6E-F, 6I**). Obviously, excess ROS, either generated from mitochondria or from peroxisomal β-oxidation of VLCFAs, can be detrimental and cause cellular stress ^64^. Notably, neurons that are more active have been documented to generate more peroxidated FAs to mitigate ROS ^62,71,72^, which is supported by our transcriptomic data (**Fig. 4C-E, Tables S6 and S10**). However, low levels of ROS can also be beneficial, for example, by promoting longevity ^95–97^. In fact, our observed increase of lactate release as an energy source in mAst co-cultured with T/T iGlut (**Fig. 6I**) may also be partially attributed to astrocytic ROS ^69,70^. Moreover, maintaining a proper level of ROS in astrocytes may be important to excite neurons by reducing astrocytic glutamate uptake. In support of this protective role for ROS, we observed reduced glutamate uptake of mAst co-cultured with T/T iGlut through elevated neuronal CLU (**Fig. 7C**), which was reversed by ROS inhibition in mAst (**Fig. 7A and B**). Interestingly, the role of ROS appears to be conserved in fibroblasts, where it also has been demonstrated to reduce glutamate uptake ^83^. Finally, our transcriptomic data also show a remarkable upregulation of the GSH metabolism (**Fig. S8A**) that can neutralize ROS ^84^, suggesting a possible ROS-feedback mechanism in LD accumulated mAst to maintain ROS homeostasis. Therefore, the AD protective allele of *CLU* and the elevated neuronal CLU may manifest beneficial effects in multiple ways involving lipid-mediated neuron-glia communication (**Fig. 7G**).

Our study using the iPSC-derived neurons co-cultured with mAst as a cellular model provides *in vitro* evidence of the protective effects of rs1582273 at the *CLU* locus, though these findings warrant confirmation using *in vivo* models. Our iPSC-derived neurons co-cultured with mAst, in combination with CRISPR/Cas9-SNP editing, provide a unique tractable model that enables us to reliably tie the AD protective allele to its neuron-specific function in mediating neuron-glia communication. Furthermore, the observed effects of the AD protective allele on neuronal CLU expression are also supported by higher expression of CLU in the brains of non-AD individuals (**Fig. 1M**), and the observed protective effect of CLU on neuronal excitation is consistent with a previous study using a mouse model ^29^. We provide strong supportive evidence from our assays based on neuronal network activity, astrocytic LD accumulation, and ROS production regarding the beneficial neuronal excitation promoted by the AD protective allele. In sum, our results provide a mechanistic link between the AD protective allele and a previously unappreciated role of neuronal CLU in mediating astrocytic LD accumulation and ROS production. This study underscores the importance of CLU-mediated complex interplay between this genetic risk factor, lipid metabolism, and neuron-glia communication in AD pathology. A better understanding of these mechanisms may offer insights into targeted therapeutic approaches for AD and other neurodegenerative conditions.

## Data Availability

All data produced in the present study are available upon reasonable request to the authors

https://zenodo.org/records/11568090

## Acknowledgments

We thank the NIH NeuroBioBank for providing postmortem human brain tissue from the Harvard Brain Tissue Resource Center and the University of Miami Brain Endowment Bank. This work was supported by National Institute on Aging grants R01AG063175 (J.D. and G.T.), R01AG081374 (J.D.), RF1AG079141 (G.T.), R01 AG073260 (H.J.B.), and U01 AG072439 (H.J.B. and Josh Shulman). A.S. and L.D.G. are supported by BrightFocus fellowships. The work is also partially supported by National Institute of Mental Health grants R01MH116281 and R01MH106575 (J.D.).

## Author contributions

X.Z. designed and performed experiments and wrote the manuscript. Y.L. analyzed RNA-seq data, calcium imaging, and wrote the manuscript. S.Z. analyzed calcium imaging, RNA-seq, and ATAC-seq data, and wrote the manuscript. A.S. contributed to experiments with human brain samples. H.Z. and A.K. performed iPSC differentiation and helped with neuron differentiation. M.J.M. and L.D.G. helped with data interpretation and edited the manuscript. Z.P.P. helped with data interpretation and neuron differentiation. A.R.S., H.J.B., and G.T. helped with data interpretation and wrote the manuscript. J.D. designed and supervised the study and wrote the manuscript.

## Supplemental information titles and legends: Figures S1-S8 and Tables S1-S15

**Figure S1:**
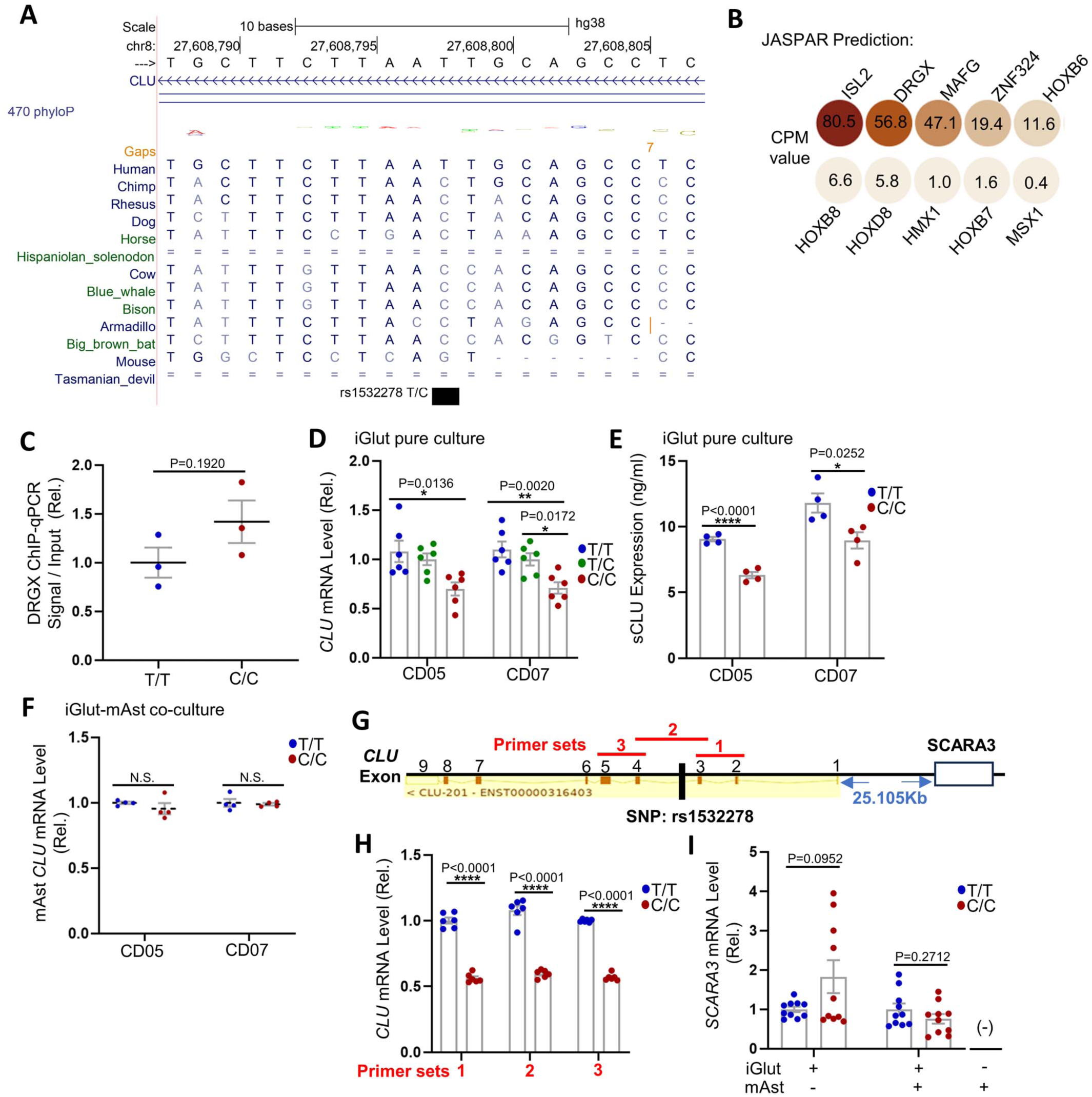
Bioinformatic and experimental validation of the regulatory effect of rs1532278 on TF-binding and *CLU* expression, related to Figure 1. (A) Multiz alignment and phyloP conservation (470 mammals) around rs1532278 (from UCSC hg38 genome browser). (B) JASPAR predicted TF binding sites at rs1532278 and TF expression levels in iGlut (10/23 predicted TFs can be detected by RNA seq). CPM, counts per million reads. (C) DRGX ChIP-qPCR for iGlut-mAst co-cultures of CD07 line on day 30. n=3 per group (one clone with 3 replicates from the CD07 line). (D) *CLU* mRNA levels in iGlut pure cultures. n=6 per group (2-3 clones per line and 2-3 replicates for each clone). (E) sCLU levels detected by ELISA from the supernatant of iGlut pure cultures. n=4 per group (2 clones per line and 2 replicates for each clone). (F) *CLU* mRNA levels of mAst in iGlut-mAst co-cultures. n=4 per group (2 clones per line and 2 replicates for each clone). (G) Three qPCR primer sets that can capture the majority of *CLU* transcript isoforms (14/17) are used in (H) to detect *CLU* mRNA levels. Only iGlut-mAst co-cultures of the CD07 line was used. n=6 per group (3 replicates per clone for 2 clones). (I) rs1532278 did not alter the expression of *SCARA3*, a *CLU*-adjacent gene, in iGlut (or when co-cultured with mAst) of the isogenic pairs of both CD05 and CD07 lines. n=10 per group (2 replicates per clone and 2-3 clones per line). Data = mean±SEM. * *p*<0.05, ** *p*< 0.01, *** *p*< 0.001, and **** *p*< 0.0001.

**Figure S2.**
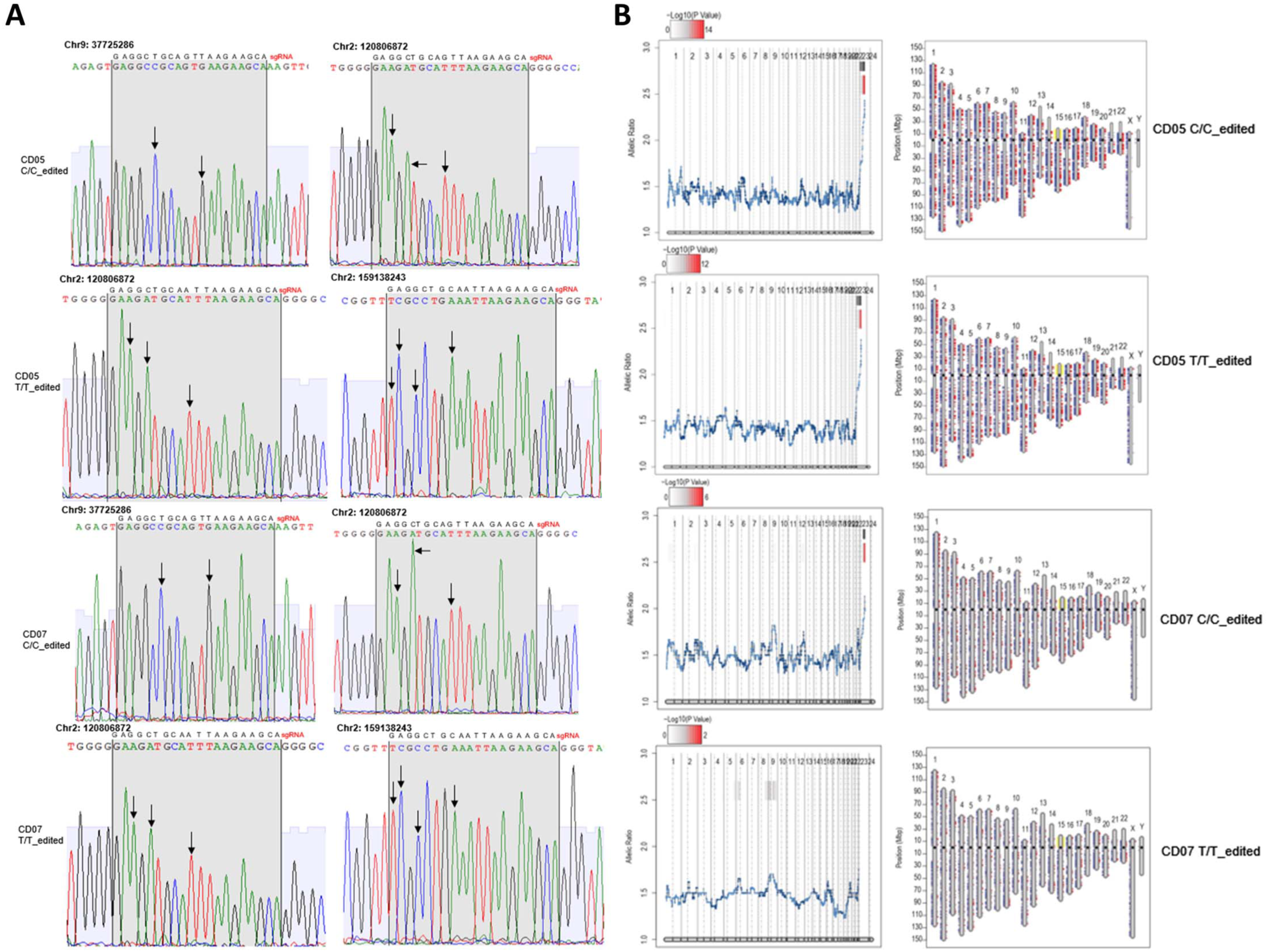
Quality control of the CRISPR/Cas9-edited iPSC lines, related to Figure 1. (A) Sanger sequencing of top off-target sites (Benching prediction) with 2 and 3 mismatched sites with gRNA in one clone of the edited alleles in both CD05 and CD07 lines. Note no off-target editing was found. (B) eSNP-karyotyping using RNA-seq data of iGlut of the two isogenic pairs of CRISPR-edited lines (CD05 and CD07; only the iPSC clones used for cellular functional assays were analyzed). Left panel, moving average of the SNP allelic ratio (RNA-seq reads of each allele) along the genome; right panel, stretches of SNP heterozygosity of all common SNPs for each chromosome. Note that no chromosomal abnormalities were found.

**Figure S3.**
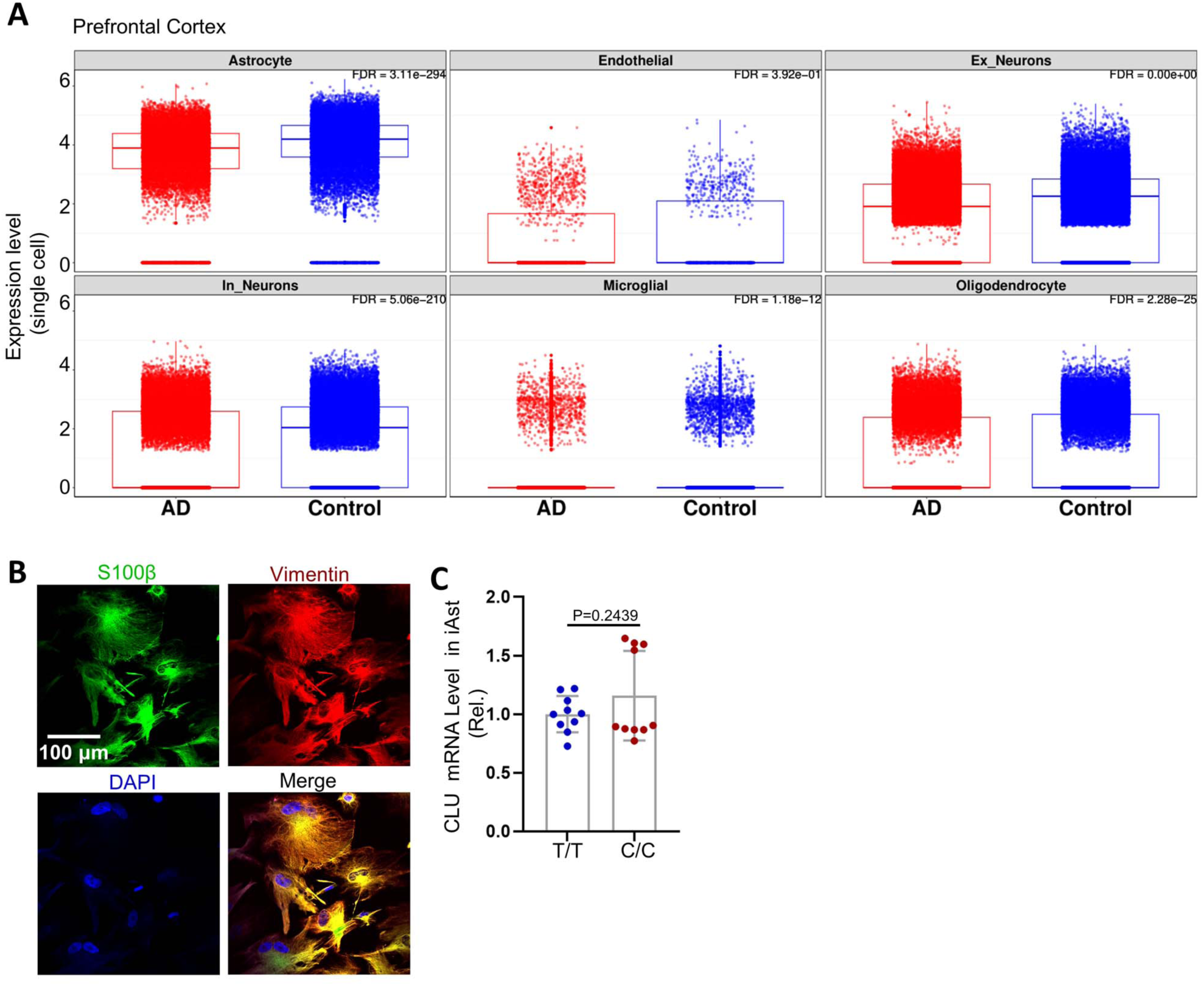
*CLU* expression in different cell types of postmortem brains of AD patients and controls, related to Figure 1. (A) Differential *CLU* expression between AD patients and controls in different cell types from data analysis of scRNA-seq data sets of the prefrontal cortex ^51,52^. Ex_neuron: excitatory neurons; In_neuron: inhibitory neurons. (B-C) CLU mRNA Levels in iAst differentiated from the isogenic pairs of CRSIPR/Cas9-edited iPSC lines (for donor lines CD05 and CD07). Immunofluorescence staining of S100β and vimentin shows the identity and purity of astrocytes. n=10 per group (2-3 clones per line and 2 replicates per clone). * *p*<0.05, ** *p*< 0.01, *** *p*< 0.001, and **** *p*< 0.0001.

**Figure S4. Synaptic and electrophysiological properties of iGlut carrying TT or C/C alleles of rs1532278 at the *CLU* locus, related to Figure 2**

(A) Quantification of SYP and PSD-95 puncta density. n=15-17 neurons per group (1-2 neurons per coverslips, 5-6 coverslips per clone, and 2 clones per line). (B-C) Western blotting shows PSD-95 and SYP levels in day-30 iGlut pure cultures. n=4 samples per group (2 clones per line and 2 biological replicates per clone). (D-E) Number of bursts and synchronicity analysis in MEA. AUNCC: Area Under Normalization Cross-Correlation. n=9-12 wells per group (2-3 clones per line and 3-4 wells per clone). Data, mean±SEM. * *p*<0.05, ** *p*< 0.01, *** *p*< 0.001, and **** *p*< 0.0001.

**Figure S5.**
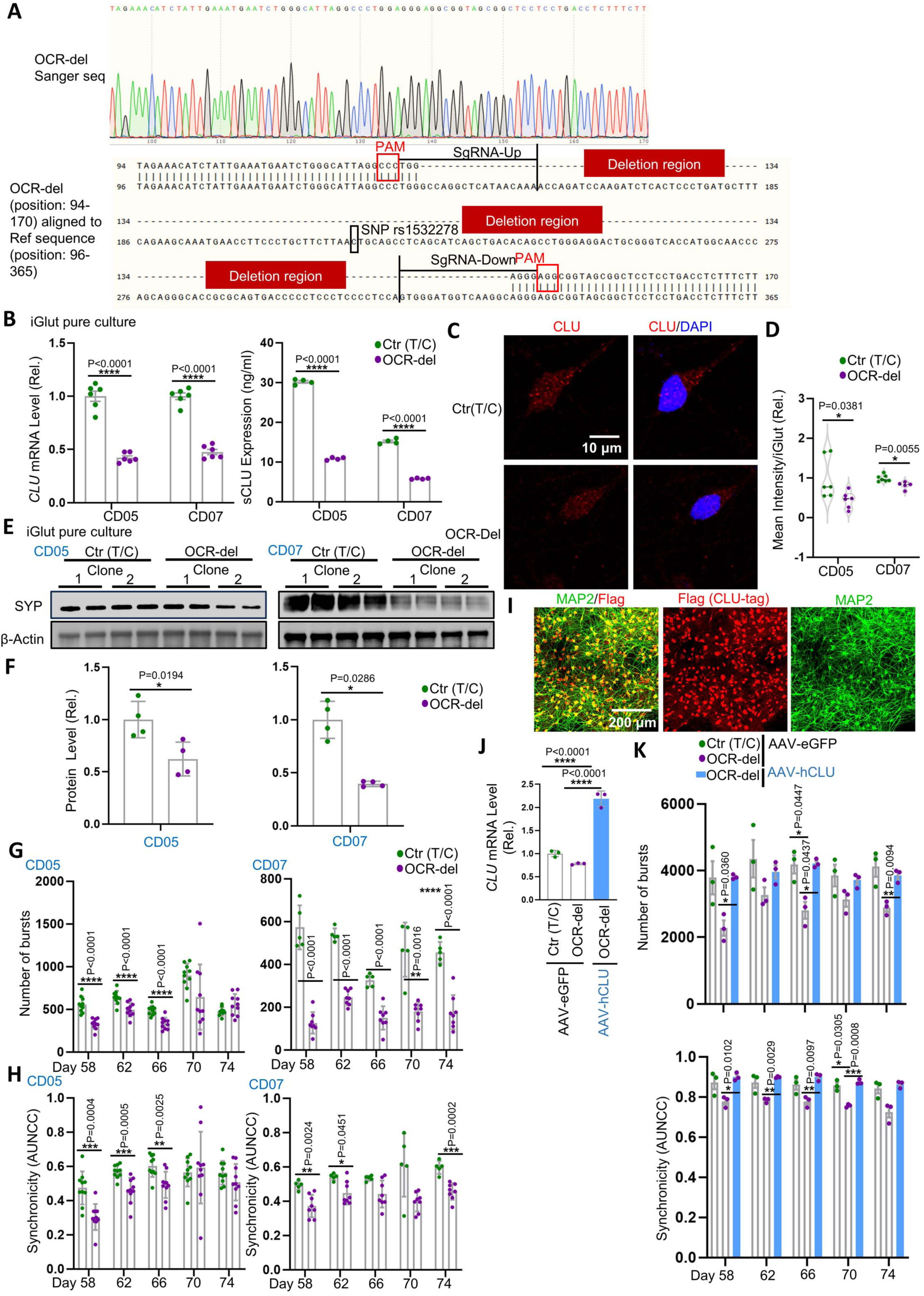
Characterization of neurons carrying the rs1532278-flanking OCR deletion or with exogenous *CLU* overexpression, related to Figure 3. (A) Sanger sequencing traces the CRISPR/Cas9-engineered homozygous OCR deletion (representative result of the CD07 line). (B) *CLU* mRNA levels (n=6 per group, 2 clones per line, and 3 replicates for each clone) and sCLU levels (n=4 per group, 2 clones per line, and 2 replicates for each clone) in iGlut pure co-cultures. (C) Immunofluorescence staining of neuronal CLU in iGlut pure cultures on days 23-25. (D) Quantification of CLU staining shown in panel (C). n=5-7 coverslips per group from two independent experiments (2 clones per line, 2-3 coverslips for each clone, and 4-5 cells averaged on each coverslip). (E-F) Western-blot for SYP in day-30 iGlut-mAst co-cultures. n=4 per group. (G-H) Number of neuron network bursts and synchronicity analysis in MEA. AUNCC: Area Under Normalization Cross-Correlation. n=5-10 wells per group (2 clones per line and 2-5 wells per clone). (I) Immunofluorescence staining of MAP2 and CLU (Flag tag) to determine AAV-hCLU transduction efficiency (∼100%). (J) qPCR to confirm *CLU* overexpression. (H) Number of network bursts and synchronicity analysis in MEA. AUNCC: Area Under Normalization Cross-Correlation. n=3 wells per group (3 replicated wells, all from one clone of line CD07). Violin plots showed data with median and interquartile range, and all other statistical graphs showed data with mean±SEM. Scale bars are indicated in corresponding images. * *p*<0.05, ** *p*< 0.01, *** *p*< 0.001, and **** *p*< 0.0001.

**Figure S6.**
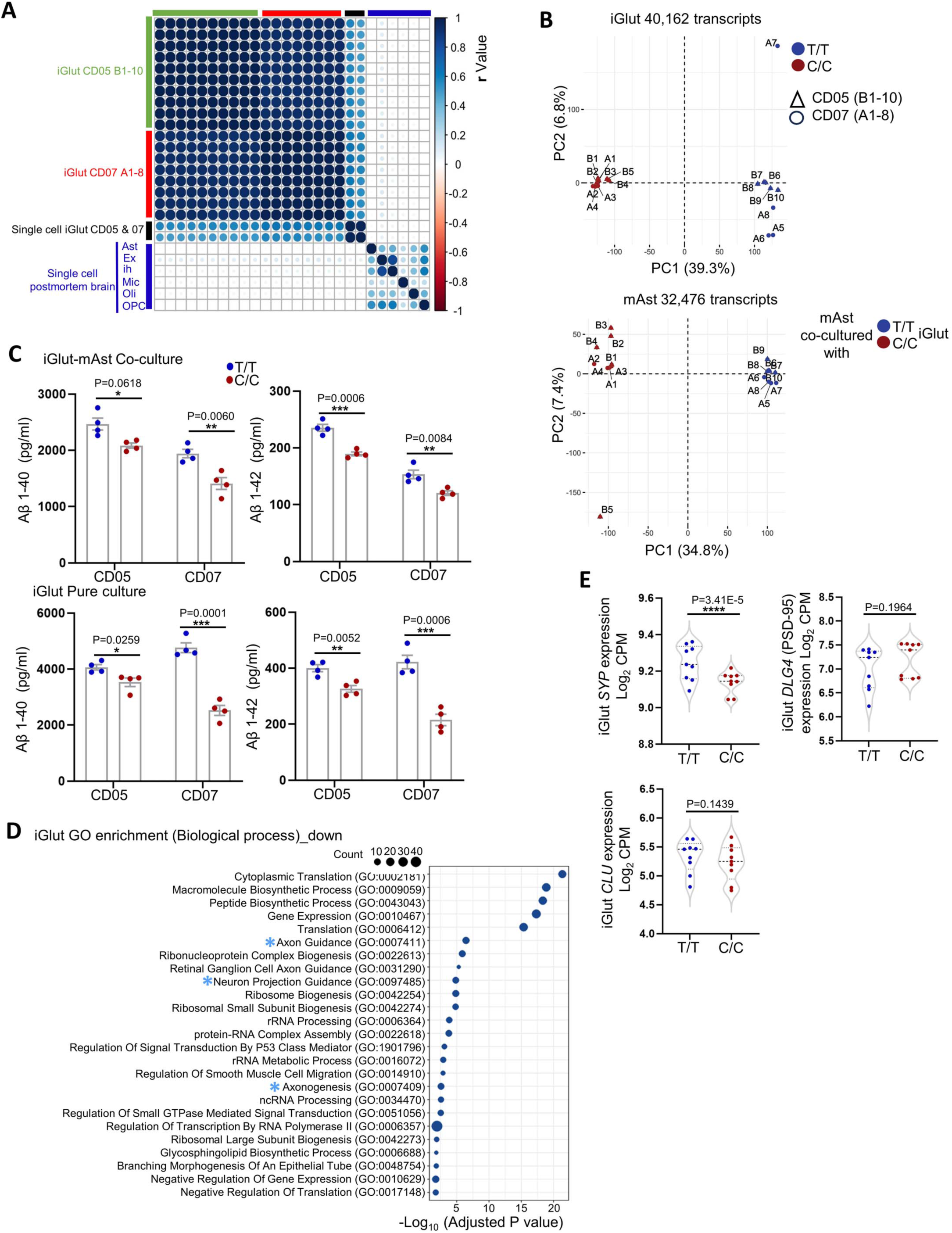
Transcriptomic analysis of iGlut-mAst co-cultures and validation, related to Figure 4. (A) Correlation analysis Salmon sorted iGlut data set to our previous published single-cell data sets iGlut (CD05 and CD07) and others published single cell data sets in postmortem brain with various cell types including ex-neuron, ih-neuron, microglia, astrocyte, oligodendrocytes, and OPC. Ast: astrocyte, ex: excitatory neuron, ih: inhibitory neuron, mic, microglia, Oli: oligodendrocyte, and OPC: oligodendrocyte progenitor cell. (B) MDS (Multidimensional scaling) analysis of all transcripts in iGlut and mAst, which are expressed in ≥75% of samples. (C) ELISA quantification of Aβ in the supernatants of iGlut-mAst co-cultures and iGlut pure cultures. n=4 per group (2 clones per line and 2 replicates for each clone). (D) GO Biological process enrichment in significantly downregulated gene sets in iGlut. Blue stars indicate neuron morphology-related pathways. (E) Confirmation of *SYP*, *PSD-95*, and *CLU* expression levels in iGlut in RNA seq analysis. * *p*<0.05, ** *p*< 0.01, *** *p*< 0.001, and **** *p*< 0.0001.

**Figure S7.**
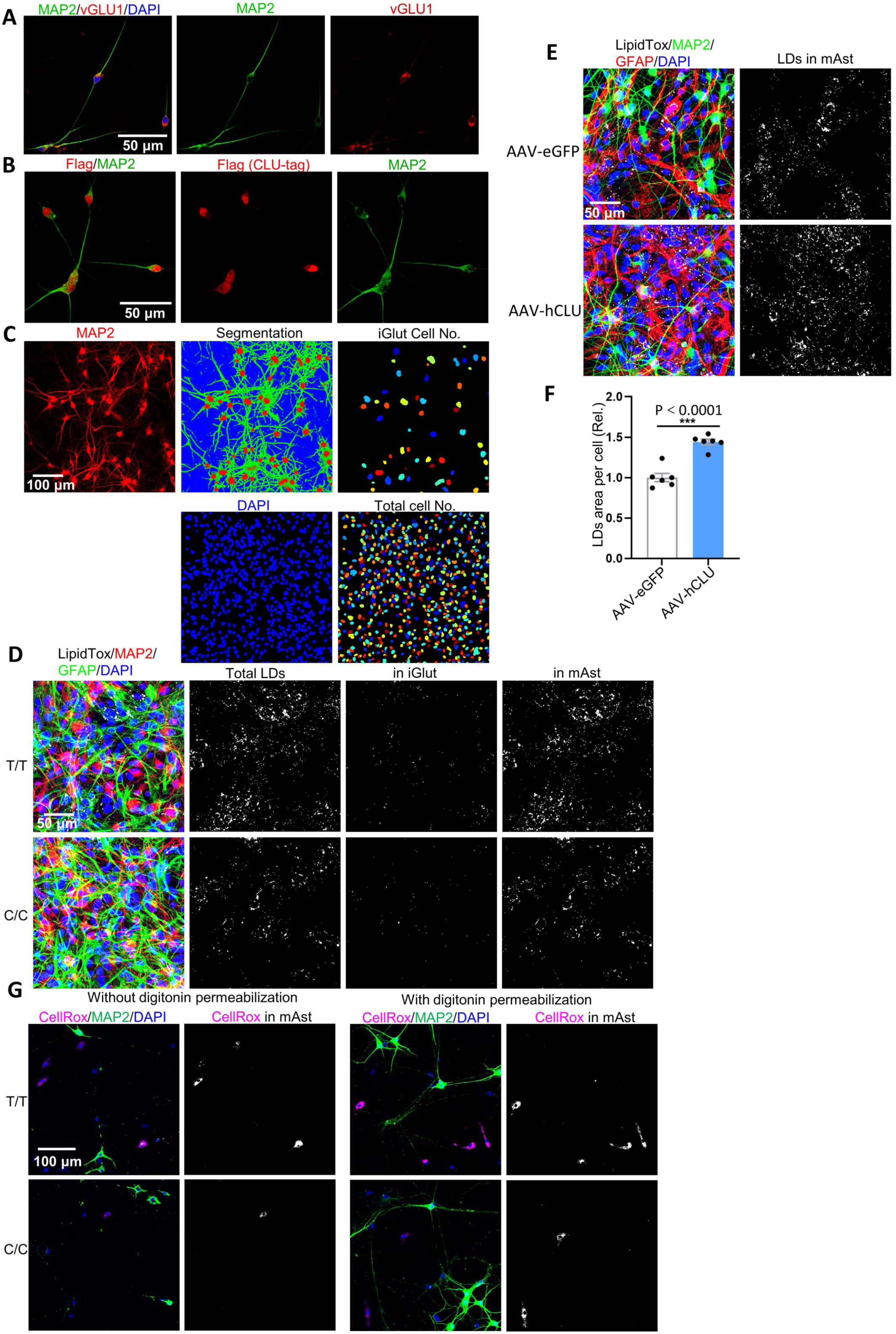
Neuronal CLU facilitates lipid transfer to astrocytes and LD accumulation accompanied by ROS production, related to Figures 5 and 6. (A) Morphology of day 23-25 iGlut pure cultures. (B) Confirmation of successful AAV-hCLU infection on day 9 in iGlut pure cultures (day 24-25) stained with Flag (CLU-Flag) and MAP2. (C) The process of cell segmentation in iGlut-mAst co-cultures, related to Fig. 5A. (D) Images from CD05 co-cultures for LDs staining, related to Fig. 6B-C. (E-F) LDs of mAst in day-30 iGlut-mAst co-culture after CLU overexpression. LDs, LipidTox+; neurons, MAP2+; astrocytes, GFAP+. n=6 coverslips per group (3 FOV averaged from each coverslip). (G) Digitonin permeabilization (1:20) does not affect CellROX staining (representative images from CD05 co-cultures).

**Figure S8.**
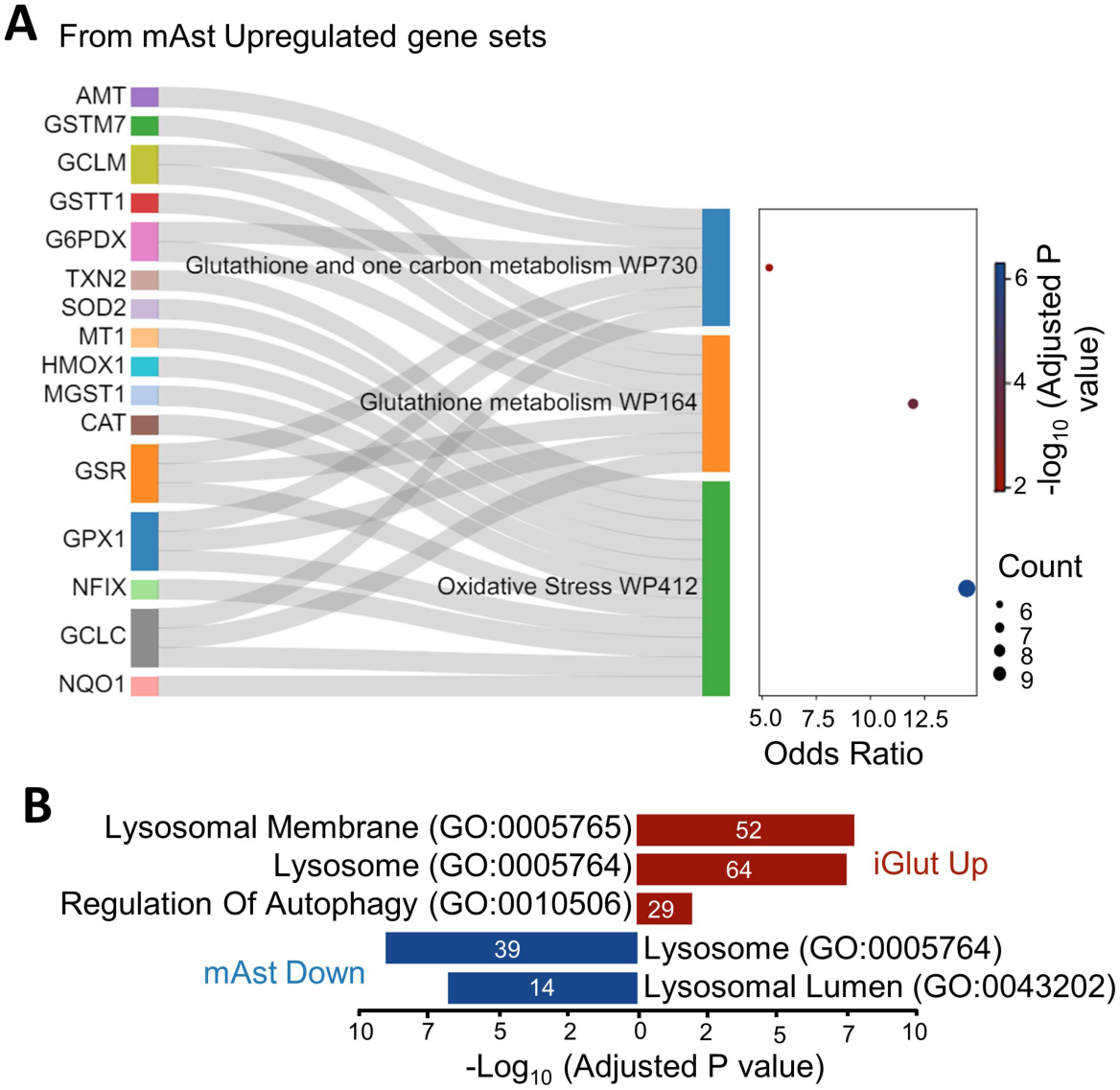
Transcriptomic data provide mechanistic support to lipid accumulation and ROS homeostasis in mAst, related to Figures 6 and 7. (A) A Sankey plot depicts glutathione metabolism and oxidative stress related pathways in mAst from Wiki pathway analysis (B) Lysosome and autophagy related GO items are enriched in DE gene lists of iGlut and mAst.

**Table S1.** ASoC SNPs that are in linkage disequilibrium with AD GWAS index risk SNP rs11787077 at CLU locus, related to Figure 1.

**Table S2.** Brain eQTL SNPs for CLU, related to Figures 1 and S3.

**Table S3.** Transcriptomic correlation between different cell types, related to Figure S6A.

**Table S4.** Differentially expressed genes in iGlut (T/T vs. C/C lines), related to Figure 4B.

**Table S5.** Differentially expressed genes in mAst (co-cultured with T/T vs. C/C iGlut), related to Figure 4B.

**Table S6.** Upregulated WikiPathways in iGlut (T/T vs. C/C lines), related to Figures 4C and 4D.

**Table S7.** Downregulated WikiPathways in iGlut (T/T vs. C/C lines), related to Figure 4.

**Table S8.** Upregulated WikiPathways in mAst (co-cultured with T/T vs. C/C iGlut), related to Figure 4F.

**Table S9.** Downregulated WikiPathways in mAst (co-cultured with T/T vs. C/C iGlut), related to Figure 4.

**Table S10.** Upregulated GO terms (biological process) in iGlut (T/T vs. C/C lines), related to Figure 4E.

**Table S11.** Downregulated GO terms (biological process) in iGlut (T/T vs. C/C lines), related to Figure S6D.

**Table S12.** Upregulated GO terms (biological process) in mAst (co-cultured with T/T vs. C/C iGlut), related to Figure 4G.

**Table S13.** Upregulated GO terms (cellular component) in iGlut (T/T vs. C/C lines), related to Figure S8A.

**Table S14.** Downregulated GO terms (cellular component) in mAst (co-cultured with T/T vs. C/C iGlut), related to Figure S8A.

**Table S15.** gRNA sequences, qPCR primers and AAV plasmid construct information, related to STAR Method.

## STAR Method

### RESOURCE AVAILABILITY

#### Lead contact

Further information and requests for resources and reagents should be directed to the lead contact, Dr. Jubao Duan.

#### Materials availability

This study did not generate new unique reagents. The iPSC lines used in this study, including the ones derived from CRISPR/Cas9 editing, are available upon request.

#### Data and code availability

iGlut-mAst co-culture RNA-seq data are available in Gene Expression Omnibus under accession code GSE269153. Two sets of ATAC-seq data for iMG, iAst, iGlut, iGABA and iDA neurons are accessible at Gene Expression Omnibus under accession code GSE263804 and GSE188941. All codes used in the analyses are accessible at https://zenodo.org/records/11568090.

### EXPERIMENTAL MODEL AND SUBJECT DETAILS

#### iPSC generation and maintenance

Two human iPSC lines, CD05 and CD07 (full IDs: CD0000005 and CD0000007), were used for CRISPR/Cas editing. These two iPSC lines were initially derived from cryopreserved lymphocytes (CPLs) using the genome-integration-free Sendai virus method (Cytotune Sendai Virus 2.0; Invitrogen) at the Rutgers University Cell and DNA Repository (RUCDR) (also known as NIMH Stem Cell Center and Infinity Biologix, currently Sampled). The two iPSC lines have been used in our previous ASoC mapping studies ^41–43^. The two donors are all healthy control males of European ancestry aged 65 and 59 for lines CD05 and CD07, respectively. The donors were also analysed for copy number variants (CNVs), and none have large CNVs (>100 kb) ^98^. They were heterozygous (T/C) at SNP site rs1532278, and the two iPSC lines were CRISPR/Cas9-edited to homozygous T/T and C/C. Two to three clones per genotype were obtained. Quality control measures for the unedited and edited iPSC lines included IF staining for pluripotency, mycoplasma contamination test, RNA-seq-based pluripotency test (Pluritest), and eSNP-karyotyping as previously described ^41,42^. The NorthShore University HealthSystem institutional review board (IRB) approved this study.

For iPSC maintenance, cells were cultured using mTeSR Plus media (StemCell Technologies, #100-0276), with medium changes performed every other day, and passaged as clumps every 4-6 days using ReLeSR (StemCell Technologies, #100-0483).

#### CRISPR-cas9 editing and Sanger sequencing confirmation

Online tool Benchling (https://www.benchling.com/) was used to design CRISPR guide RNA (gRNA), and the gRNAs with highest off-target score (specificity) were selected (**Table S15**). The gRNAs were cloned into CROPseq-Guide-Puro vector (#86708, Addgene) ^99^. The gRNA plasmid DNAs, together with the plasmid DNAs of pSpCas9(BB)-2A-Puro (#62988, Addgene)^100^ and pSUPERIOR.puro-shp53 (#38035, Addgene) (included to transiently inhibit p53 and increase editing efficiency ^101^) were transiently transfected into iPSCs. For transfection, iPSCs were dissociated into single cells using accutase (07920, StemCell) in a 15ml centrifuge tube at a density of 4-6 × 10^5^/1.8ml in the presence of 10 μM ROCK inhibitor (ROCKi, 1254/1, R&D Systems). Lipofectamine stem reagent (STEM00001, Thermo Fisher Scientific) was used for transfections. DNA including pSpCas9(BB)-2A-Puro (0.5 μl from 1 μl/ug stock), CROPseq-Guide-Puro vector (1 μl from 1μl/ug stock), pSUPERIOR.puro-shp53 vector (1 μl from 1μl/ug stock) and ssODNs carrying the desired SNP allele (6 μl from 100uM stock) were mixed into 100 μl Opti-MEM media (31985062, Thermo Fisher Scientific) in a 1.5ml tube. Another 1.5 ml tube was filled with 8 μl Lipofectamine stem reagent and 100 μl Opti-MEM media. The reagents in the two 1.5 ml tubes were combined and added into iPSC single cell suspension, followed by gently mixing and replating into one well of a 6-well plate. Sixteen hours post-transfection, the medium was replaced with fresh medium containing 0.25 μg/ml puromycin (A1113802, Thermo Fisher Scientific) and 10 μM ROCKi. Forty-eight hours post transfection, the medium was refreshed with the same reagents. Sevety-two hours post transfection, the puromycin-containing medium were removed, and the cells were cultured in fresh medium with 5 μM ROCKi. Half-medium (without ROCKi) change was made every 2-3 days until iPSC colonies were formed. 10-14 days post transfection, iPSC colonies were individually picked into 96-well plates. DNA from a small fraction of cells in each colony was then extracted (QE09050, Epicentre) and used for Sanger sequencing confirmation of accurate editing. Subcloning with at a low density (2-5000 cells on 6-cm dish) was carried out to ensure an iPSC clone is pure. Following successful editing, three predicted top-ranking off-target editing sites were subjected to Sanger sequencing to confirm the absence of off-target editing (**Table S15 and Fig. S2A**).

For OCR deletion, up- or downstream gRNAs were cloned into the vector pSpCas9(BB)-2A-Puro to co-express gRNAs and Cas9. Transfection was conducted as described above. The presence of on-target OCR deletion and the absence of the predicted off-target editing in the selected iPSC clones were confirmed by Sanger sequencing.

#### Preparation of primary mouse astrocytes

Primary mouse astrocytes were extracted as previously described ^102^. Briefly, the brains were harvested from Day 0-2 pups of C57BL/6J. Their meninges were removed, minced and centrifugated in cold Hanks’ balanced salt solution (HBSS buffer, 88284, Thermo Fisher Scientific). Then, tissue pellets were resuspended, dissociated and filtered through a cell strainer to generate a single cell suspension. All cells were seeded into T-75 flasks, maintained in DMEM (10569016, Thermo Fisher Scientific) with 10% FBS (A5209501, Thermo Fisher Scientific). The obtained astrocytes were used within a month (≤4 cell passages).

#### Glutaminergic neuronal (iGlut) differentiation

iGlut neurons were differentiated from iPSC according to the previous protocols ^42,103^ with some modifications. Briefly, on day −1, iPSCs were dissociated with accutase and placed in 6-well plates at a density of 5 × 10^5^/well in the presence of 5 μM ROCKi. On day 0, NGN2 and rtTA lentivirus were added into mTeSR Plus medium with 5 μM ROCKi to infect iPSCs. On day 1, Neurobasal media (21103049, Thermo Fisher Scientific) containing 1× B27 supplement (17504044, Thermo Fisher Scientific), 1x GlutaMax (35050061, Thermo Fisher Scientific), 5 μg/ml Doxycycline (Dox, D9891, Sigma) and 5 μM ROCKi were used to initiate the differentiation. 1μg/ml puromycin selection was performed from Day 2 to Day 4. On day 5, the cells were dissociated with accutase, replated into matrigel-coated plates, and maintained in Neurobasal media supplemented with 1× Glutamax, 1x B27, 5 µg/ml doxycycline, 10 ng/ml BDNF/GDNF/NT-3 (450-02/450-10/450-3, Pepro Tech), and 5% FBS. On day 6, the culture medium was refreshed with the same one used on Day 5 without FBS, and 1 µM Ara-C (C6645, Millipore-Sigma) was applied to the medium and kept for two days to ensure the purity of postmitotic neurons. On day 8, Ara-C was withdrawn and the neurons were maintained with one half-medium change every 3 days until they were mature enough for the assays.

For CLU and LDs staining in pure iGlut culture, day-5 neurons were directly replated onto 12 mm glass coverslips (GG-12-15-Pre, Neuvitro) at 50,000 cells/coverslip. Neurons were maintained on the coverslip until day 25 by refreshing half the medium every three days. For neuron morphology and LD staining in the iGlut-mAst co-culture system, 100,000 astrocytes were first placed onto glass coverslips 24 hours before dissociating neurons in 24-well plates with DMEM medium with 10% FBS. Then, day-8 iGlut neurons were dissociated with accutase, suspended in Neurobasal medium, and seeded on top of the layer of astrocytes at 300,000 cells/well after removing the DMEM medium. On day 9, one-half of the medium was replaced without FBS, and on day 10, all medium were refreshed to remove FBS completely. Thereafter, all medium was refreshed every 3 days until day 30. For ChIP-qPCR and RNA seq purposes, day-8 iGlut and mAst were dissociated and placed together at appropriate dishes/plates, and the cultures were maintained the same way as iGlu-mAst co-culture described above.

#### iPSC differentiation into astrocytes (iAst)

iAst were differentiated from iPSC based on the NgN2 method ^53^ with minor modifications. Briefly, on day −1 and day 0, the same iGlut differentiation procedures were followed. On day 2, a cocktail of SB431542 (10 µM) (1614, Tocris Bioscience), XAV939 (2 µM) (3748, Tocris Bioscience), LDN193189 (100 nM) (6053, Tocris Bioscience), Dox (5 µg/ml) and ROCKi (5 µM) were added into N2 medium that was constituted with DMEM/F12 (11320-033, Gibco), 1x Glutamax, 0.3% Sucrose (S0389, Millipore-Sigma), and 1x N2 supplement B (07156, StemCell Technologies). On day 3, SB431542 (5 µM), XAV939 (1 µM), LDN193189 (50 nM), Dox (5 µg/ml) and Puromycin (5 µg/ml) were added into N2 medium to start cell selection. On day 4, the culture medium was refreshed with N2 medium supplemented with Dox (5 µg/ml) and Puromycin (5 µg/ml). On day 5, all cells were dissociated with Accutase, suspended with Astrocyte medium (1801, SiceneCell) containing 5 µM ROCKi, and replated onto Matrigel-coated 10-cm dishes. Cells were passaged at 1:2 ratio in Astrocyte medium every 3-4 days. On day 25, the cells were fixed with 4% PFA, permeabilizated with 0.1% Triton X-100 (SLCD3244, Sigma) and stained with S100β (S2532, Millipore-Sigma, Mouse, 1:500) and Vimentin (D21H3, Cell Signaling, Rabbit, 1:200) to determine iAst purity (∼100%).

#### iGlut morphology and CLU staining

Day-30 iGlut-mAst co-culture on coverslips were fixed with 4% PFA for 15 min and permeabilized with 0.1% Triton X-100 for 20 min. The fixed cells were incubated with antibodies again PSD-95 (clone K28/43, NeuroMab, Mouse, 1:1,000), SYP (ab32127, Abcam, Rabbit, 1:500), and MAP2 (188004, Synaptic System, Guinea pig, 1: 1000) overnight at 4°C. After three times of washing with PBS, the corresponding secondary antibodies including Donkey anti-Mouse Alexa 488 (A21202, Thermo Fisher Scientific, 1:200), Donkey anti-Rabbit Alexa 594 (A21207, Thermo Fisher Scientific, 1:200), and Goat anti-Guinea Pig Alexa 647 (A21450, Thermo Fisher Scientific, 1:200) were added and further incubated for two hours at room temperature. Nuclei were stained with DAPI (0.5 mg/ml) for two minutes at room temperature before the coverslips were mounted (Fluorescent Mounting Medium, F4680, Millipore-Sigma). For IF staining of neuron identity, the cultures were stained with HuNu (MAB1281, Millipore-Sigma, Mouse, 1:200) and MAP2 antibodies. All antibodies were dissolved in PBS with 1% BSA (15260037, Thermo Fisher Scientific) and 0.1% Triton X-100.

For CLU staining in pure culture iGlut, cells were fixed on day 25, permeabilized, and incubated with CLU (ab69644, Abcam, Rabbit, 1:300) and NeuN (MAB377, Millipore-Sigma, Mouse, 1:100) antibodies. NeuN immunostaining was used to visualize perikaryon where most CLU signals were located (in the **Image quantification** section below). All procedures were the same as above. In addition, immunostaining of vGlut1 (ab227805, Abcam, Rabbit, 1:500) and MAP2 were used to determine the purity of iGlut cultures.

#### RNA isolation from cell cultures for qPCR and RNA-seq

Total RNA was extracted using the RNeasy Plus Kit (74134, Qiagen). Briefly, all cells were directly lysed in RLT plus buffer, further separated and eluted in RNase-free water. For RNA-seq, all RNA samples were sequenced on the Illumina NovaSeq 2000 platform with paired-end reads (2 × 150 bp) (Novogene). For qPCR, RNAs were first reversed transcribed to cDNAs using a high-capacity cDNA reverse transcription kit (4368814, Applied Biosystems) and further amplified with TaqMan Universal PCR Master Mix (4364338, Applied Biosystems) on a Roche 480 II instrument. All qPCR primers were listed in **Table S15**.

#### Chromatin immunoprecipitation (ChIP) qPCR

ChIP-qPCR assay was performed following the protocol of Magna ChIP A/G Chromatin Immunoprecipitation Kit (17-10085, Millipore-Sigma). Briefly, 1% formaldehyde (28908, Thermo Fisher Scientific) was directly added into the medium of day-30 iGlut-mAst co-cultures to cross-linking proteins to DNA. Then, glycine was added to terminate the cross-linking. Cells were harvested in ice-cold PBS, pelleted, sonicated, and immunoprecipitated with 10 µg ISL2 (AF4244, R and D Systems, Sheep) or DRGX antibodies (bs-11827R, Bioss, Rabbit) and Protein A/G magnetic beads for overnight at 4°C. Protein A/G magnetic beads with TF-binding DNAs were separated on a magnetic stand. TF-DNA complexes were eluted in an elution buffer with proteinase K at 62°C with 2 hours of shaking, followed by a 95°C incubation for 10 minutes to denature proteinase K. Lastly, the released DNAs were purified for qPCR. About 2% of sample input after sonication without immunoprecipitation was used to normalize the loading input. Normal sheep IgG (5-001-A, R&D Systems) or rabbit IgG (2729, Cell Signaling Technology) were used as negative control to exclude non-specific binding of antibodies. The ChIP-qPCR primers are listed in **Table S15**.

#### *ISL2* siRNA knockdown

iGlut were directly replated onto a 24-well plate at 500,000 cells/well on Day 5. On day 30, the neurons were transfected either by 50 nM human I*SL2* siRNA (L-016725-00-0005, Horizon Discovery Biosciences) or 50 nM non-targeting control siRNA (D-001810-10-05, Horizon Discovery Biosciences) following the protocol of Lipofectamine RNAiMAX Transfection Reagent (13778030, Thermo Fisher Scientific). 72 hours post transfection, RNAs were extracted to quantify *ISL2* and *CLU* mRNA levels by qPCR. The qPCR primers are listed in **Table S15**.

#### RNA isolation from postmortem brain samples and qPCR

Frozen human brain samples (frontal pole BA10 region) were received through the NIH biobank from Harvard Brain Tissue Resource Center and the University of Miami Brain Endowment Bank (Reference ^46^ extended Table 10) and stored at −80°C. The grey matter was dissected from brain blocks on dry ice. RNA was isolated using a Direct-zol RNA MiniPrep Kit (R2050, Zymo), per the manufacturer’s instructions. RNAs were reverse-transcribed into cDNA using a high-capacity cDNA reverse transcription Kit, per the manufacturer’s instructions. Reactions were set up using PowerUp SYBR Green Master Mix for qPCR (4368814, Applied Biosystems) and run on a QuantStudio3 Real-Time PCR System (Applied Biosystems). The data were analyzed using 2-ΔΔCt method and normalized to *COTL1*. The primer sequences are listed in **Table S15**.

#### Enzyme-linked immunosorbent assay (ELISA) assay

For both iGlut pure culture and iGlut-mAst co-culture, the culture medium was completely refreshed on day 27 and further collected on day 30. The conditioned media were centrifuged for 10 min with 3000 rpm at 4°C to remove the cell debris. Cells were harvested for BCA protein quantification to normalize the ELISA detection from the supernatant. ELISA quantifications of human CLU (DCLU00, R&D system, specific for human CLU), human Aβ 1-40 (DAB140B, R&D system), and human Aβ 1-42 (DAB142, R&D system) were performed according to the vendors’ protocol.

#### Western Blotting

Western blotting was performed as previously described ^102^. Briefly, cells were lysed in NP-40 lysis buffer (J60766-AK, Thermo Fisher Scientific) with proteinase inhibitors (04693159001, Roche) and phosphatase inhibitors (04906845001, Roche). The cell lysates were sonicated and denatured in Laemmli Sample Buffer (1610747, Bio-Rad). BCA kit (23225, Thermo Fisher Scientific) was used for total protein quantification. The cellular protein extracts were fractioned through 10% homemade SDS-PAGE gels, transferred to a PVDF membranes (1620177, Bio-Rad) and subsequently immunoblotted. The primary antibodies included PSD-95 (ab18258, Abcam, Rabbit, 1:1,000), SYP (ab32127, Abcam, Rabbit, 1:2,000), and β-actin (3700, Cell Signaling Technology, 1:5,000). The secondary antibodies included anti-rabbit-HRP (7074, Cell Signaling Technology, 1:5,000) and anti-mouse-HRP (7076, Cell Signaling Technology, 1:5,000). All Western blotting images were quantified using FIJI ^104^, and specific protein signals were normalized to corresponding β-actin signals in each sample.

#### Calcium imaging

iGlut were infected with AAV-GCaMP6m (pAAV.Syn. GCaMP6m.WPRE.SV40, 100841-AAV9, Addgene) or AAV-jRCaMP1b (pAAV.Syn.NES-jRCaMP1b.WPRE.SV40, 100851-AAV9, Addgene) at 10^5^ MOI on day 8. The infected neurons were replated together with mAst into a 96-well plate (ANFS-0096, Curi Bio) on day 9 at a density of 15,000 iGlut and 5,000 mAst per well. On day 35, the time-lapse images were acquired at ∼5 Hz for 2 min on a Nikon A1R microscope with sCMOS camera. For quantification, peak detection was performed using the R baseline package, and HDBSCAN was used to screen out the correct clusters.

#### MEA

MEA assay was performed according to a previous protocol ^42^ with slight modification. Briefly, day-20 iGlut were disassociated with accutase and replated with mAst into 0.1% PEI-coated 24-well MEA plate (M384-tMEA-24W, Axion BioSystems) at a density of 150,000 iGlut and 50,000 mAst. Neurobasal medium with 1× Glutamax, 1x B27, 5 µg/ml doxycycline, 10 ng/ml BDNF/GDNF/NT-3, and 5% FBS were used on the first day after replating. In the next two days, half of the medium (300 µL) was replaced every day with fresh culture medium without FBS. Thereafter, 2/3 culture medium (no FBS) was regularly refreshed every 3 days. The culture medium was completely refreshed a day before the MEA recording. Spontaneous firing was recorded for 15 minutes, and the last 10 minutes of recording were used for data analysis. All data files were batch-processed using the Neural Metrics Tool (Axion Biosystems). For data analysis, the burst parameters were set at Poisson Surprise with a minimum surprise of 10 and Adaptive mode, Minimum number of spikes to 40, and Minimum of electrodes to 15%; Active Electrode Criterion was set at 6 spikes/min; Synchrony parameters were set at 20 ms for Synchrony Window and “none” for Synchrony Metrics.

#### AAV plasmid reconstruction, packaging and infection

To express human *CLU*, pAAV-hSyn-hCLU-Flag was reconstructed from pAAV-hSyn-eGFP (58867, Addgene). First, pAAV-hSyn-eGFP was digested with BamHI and EcoRI to remove the eGFP insert. Then, the backbone DNAs (30 fmol), *CLU* cDNA fragment (1400 bp; 60 fmol, synthesized from IDT, **Table S15**), ssDNA (single strand)-up-linker 60 bp (200 fmol, synthesized from IDT, **Table S15**), and ssDNA -down-linker 60 bp (200 fmol, synthesized from IDT, **Table S15**) were assembled using NEBuilder® HiFi DNA Assembly Master Mix (E2621S, New England Biolabs). The assembly reaction was then transformed into NEB stable competent *E coli* (C3040H, New England Biolabs). The transformed bacterial clone with the correct insert was determined by Sanger sequencing of plasmid DNAs, and a correct clone was expanded to prepare the plasmid DNAs for AAV packaging.

Recombinant AAVs were packaged in HEK 293T cells (CRL-3216, ATCC) following a previous protocol ^105^ with minor modifications. HEK 293T cells were transfected with pAAV-hSyn-hCLU-Flag/pAAV-hSyn-eGFP, pUCmini-iCAP-PHP.eB (103005, Addgene) and pAdDeltaF6 (112867, Addgene) by PEI (23966, Polysciences) at 90% confluence. Seventy-two hours post-transfection, all cells were harvested and pelleted. AAVs were further extracted from cell pellets by AAV extraction kit (6675, Takara) and titrated using qPCR approaches (primers are listed in **Table S15**).

iGlut were infected with AAV-eGFP or AAV-hCLU at 10^5^ MOI on day 8. For calcium imaging, iGlut were co-infected with AAV-jRCaMP1b at 10^5^ MOI on the same day. On the next day, iGlut were co-cultured with mAst or alone, following the above-described iGlut differentiation procedures. Immunostaining with Anti-Flag (20543-1-AP, Proteintech, Rabbit,1:200) and MAP2 antibody was used to determine the infection efficiency of AAV-hCLU on day 30 for iGlut-mAst co-culture and on day 25 for iGlut pure culture.

#### Lipid droplet (LD) staining

For iGlut pure culture, day-25 cells were fixed with 4% PFA, washed with PBS, and stained with LipidTox (H34476, Thermo Fisher Scientific, 1:300) for 30 minutes at room temperature. LipidTox was dissolved in PBS with 1mg/ml DAPI. After staining, a one-time quick wash (∼2 seconds) with PBS was applied before mounting the slides. The images were taken immediately after mounting on a Nikon ECLIPSE TE2000-U microscope.

For iGlut-mAst co-culture, day-30 cells were fixed with 4% PFA, washed by PBS, and digitonin permeabilized (#16359, Cell Signaling Technology, 1:20) for 20 min at room temperature. Cells were then stained using antibodies against MAP2 (188004, Synaptic System, Guinea pig,1: 1000), GFAP (Z0334, DAKO, Rabbit, 1:500) and/or Anti-Flag (20543-1-AP, Proteintech, Rabbit,1:200) that were dissolved in PBS with 1% BSA. The corresponding secondary antibodies were dissolved in the same buffer to stain the cell for 2 hours at room temperature. After three times of washing by PBS, LipidTox (1:300) or BODIPY 493/503 (D3922, Thermo Fisher Scientific, 2 µg/ml) were used to stain LDs at room temperature. The staining time was 30 minutes for LipidTox and 10 minutes for Bodipy 493/503. Both dyes were dissolved in PBS with DAPI (1mg/ml). Bodipy 493/503 stocks were prepared in DMSO at 1 mg/ml. Cells were quickly washed (∼ 2 seconds) once with PBS for LipidTox staining before mounting. Three quick washes (∼2 seconds) with PBS were applied for BODIPY 493/503 staining ^74^.

#### Lipid transfer assay

The lipid transfer assay was performed as previously described ^10,62^ with minor modifications. Briefly, following the iGlut differentiation protocol, day-5 iGlut were replated into 24-well plates at 500,000/well and maintained until day 30. The mAst were replated into 24-well plates with Matrigel-precoated coverslips at 15,000/well and maintained for 3 days. Day-30 iGlut were prelabeled by 2.5 μM Red C12 (BODIPY 558/568, D3835, Thermo Fisher Scientific) by adding Red C12 directly into culture medium and incubated for 18 hours. iGlut were then washed twice with pre-warmed DPBS and rested for one hour in culture medium in a 37 °C incubator. mAst cells on coverslips and iGlut neurons in culture wells were washed twice with pre-warmed DPBS. Pre-warmed HBSS (14175095, Thermo Fisher Scientific) containing 2 mM CaCl2 and 10 mM HEPES was added to the wells with neurons. A parafilm separator, cut to suitable size and with the center removed to form a ring, was placed above the neurons. Coverslips carrying mAst were placed face-down onto the neurons, creating a sandwich structure. The assembled cultures were then incubated at 37°C for 4 hours. After incubation, coverslips with mAst were removed, fixed and stained with Bodipy 493/503.

#### Ketone body and Lactate assay

Day-5 iGlut were directly seeded into 12-transwell (3460, Corning, 0.4 μm pore) at 1 × 10^6^ cells/well and cultured until day 15. mAst were seeded into the insert of 24-transwell at 200,000 cells/insert on day 13, placed into the unused wells and rested for two days in DMEM medium with 10% FBS. Then, the culture medium in the mAst insert was replaced with the iGlut culture medium (Neurobasal medium with B27, Glutamax, BDNF, GNDF, and NT3). The mAst insert was then transferred into the wells with day-15 iGlut for 14 days of co-culture with medium change every 3 days. Then, the insert with mAst were taken out and washed three times with PBS. Then, the mAst insert was incubated in 5 times diluted DMEM medium (10569016, Thermo Fisher Scientific, supplemented with Glutmax) to reach 5mM glucose condition, without FBS and sodium pyruvate for 24 hours. The supernatants were then collected at 6 and 24 hours to measure the concentration of β-hydroxybutyrate (a ketone body, JE9500, Promega) and Lactate (J5021, Promega). iGlut and mAst were also collected with NP-40 lysis buffer to determine the protein concentration using the BCA method for normalization. Data were normalized to protein concentrations of iGlut and mAst.

#### CellRox and SLC7A11 staining

Day-8 iGlut and mAst were replated into 24-well plates with Matrigel-precoated coverslips at a density of 30,000/well for neurons and 10,000 /well for astrocytes. iGlut/mAst were maintained in the culture medium until day 30, following the iGlut-mAst coculture protocol described above. For CellRox staining, CellRox Deep Red (C10422, Thermo Fisher Scientific) was added to the culture medium (5 μM final concentration) and incubated for 30 min at 37°C. After incubation, cells were fixed with 4% PFA, and processed with and without digitonin permeabilization (**Fig. S7G**) to determine whether CellRox Deep Red signals can survive with this gentle permeabilization, and further stained with MAP2. For SLC7A11 staining, cells were fixed with digitonin permeabilization and further stained with SLC7A11 (PA116893, Thermo Fisher Scientific, Rabbit, 1:300) and MAP2.

#### Glutamate uptake assay

Day-5 iGlut were directly seeded into 24-transwell (3413, Corning, 0.4 μm pore) at 500,000 cells/well and cultured until day 15. mAst were seeded on the insert of 24-transwell at 40,000 cells/insert on day 13, placed into the unused wells and rested for two days in DMEM medium with 10% FBS. Then, the culture medium in the mAst insert was replaced with iGlut culture medium (Neurobasal medium with B27, Glutamax, BDNF, GNDF, and NT3). The mAst insert was then transferred into the wells with day-15 iGlut for 14 days of co-culture with medium change every 3 days. Then, the insert with mAst were taken out, washed once with HBSS buffer (14175095, Thermo Fisher Scientific) without calcium and magnesium, and incubated in the same buffer at 37°C for 30 min. mAst insert was incubated in HBSS buffer (14025092, Thermo Fisher Scientific) with 100 μM glutamate, calcium and magnesium for 3 hours. The supernatants were then collected to measure glutamate concentration using the glutamate assay kit (Ab83389, Abcam). iGlut and mAst were also collected with NP-40 lysis buffer to determine the protein concentration using the BCA method for normalization purpose. Data were normalized to protein concentrations of iGlut and mAst.

For antioxidant AD4 treatment, iGlu were cocultured with mAst on trans-well dishes. On day 14, all medium in cultured wells and inserts was replaced with fresh medium containing AD4 (1.5 mM final concentration) for 24 hours. Then, the inserts with mAst were removed for glutamate uptake assay as described above.

#### CLU immunodepleting assay

Day-5 iGlut were replated into 12-well at 100,000 cells/well and maintained following the protocol described above. On day 27, the medium was refreshed with iGlut culture media. On day 30, the supernatants were collected and centrifuged at 3000 rpm for 10 minutes at 4°C to remove cell debris, and then frozen and stored at −80 °C until use. CLU antibody (sc-166907, Santa Cruz) was conjugated to magnetic beads (14311D, Thermo Fisher Scientific) at 8 μg of antibody per mg of beads following the manufacturer’s protocol. 2 mg of antibody coupled with beads were added into 1 ml day-30 supernatant (16 μg antibody per ml of medium) and incubated at 4°C for 24 hours. The beads were then captured on a magnetic stand, and the supernatants were filtered through 0.2 μm syringe filters (13100106, Basix) (CLU immunodepleted medium). The conditioned medium (CM) was prepared by combining fresh DMEM medium with 10% FBS and CLU immunodepleted medium at a 1:3 ratio. For assaying the function of mAst, mAsts were seeded into 96-well plates at 40,000 cells/well 2 days before adding the CM. On day 1, the mAst culture medium was completely refreshed with CM (200 μl) and maintained until day 7 with one media change on day 4. On day 7, all CM were removed and washed with HBSS buffer (14175095, Thermo Fisher Scientific) to start the glutamate uptake assay as described above. The assay data were normalized to the protein concentration of mAst from the corresponding wells.

#### Image quantification

***Neuron morphology (MAP2 staining)***: All confocal z stacks (20x objective lens) were first processed using FIJI ^104^ to generate maximum intensity projections. Then, neuron branches and soma were segmented by ilastik ^106^. The branch length and the number of neurons (soma number) were analyzed by Cellprofiler ^107^ as previously described ^102,108^. ***SYP and PSD-95 puncta***: All confocal z stacks (63x oil immersion objective lens) were first projected with maximum intensities. ROIs of MAP2 positive dendrites were chosen uniformly from the secondary branches with ∼50-100 μm length by FIJI ^104^. The identifical fluorescence intensity thresholds were applied to different images to identify SYP and PSD95 positive puncta. SYP - and PSD95-positive objects within the MAP2 mask were analyzed. For density measurements, the number of SYP or PSD-95 objects was divided by the length of MAP2 (per unit was set as 10 μm) in an ROI; for area measurements, the total area of SYP and PSD-95 objects were divided by the area of MAP2 in an ROI. Cellprofiler was used for all quantifications. ***CLU intensity***: All confocal z stacks (63x oil immersion objective lens) were cropped into single cells, and z projected with maximum intensity in FIJI. NeuN staining was used to generate a mask of soma after setting a proper intensity threshold to encircle the majority of CLU positive signals. The mean intensity of CLU signals within the NeuN mask was quantified by Cellprofiler. ***LipidTox staining in iGlut pure culture***: All confocal z stacks (63x oil immersion objective lens) were cropped into single cell and z projected with maximum intensity by FIJI. Then, the uniform and proper threshold was set for LipidTox signals across different groups, and the area of LipidTox signals were calculated in Cellprofiler. ***Red C12 staining in mAst***: All confocal z stacks (20x objective lens) were z projected with maximum intensity in FIJI. A uniform and proper threshold was set for Red C12 signals across different groups, and the area of Red C12 signals was calculated in Cellprofiler. ***LipidTox staining in iGlut-mAst co-culture***: All channels in confocal z stacks (20x objective lens) were split by FIJI without any projection. Then, proper thresholds were set for both LipidTox and MAP2 signals. MAP2 staining was used to generate a 3D mask in Cellprofiler. LipidTox staining within MAP2 3D mask was identified as neuronal LDs, and non-overlapping staining was considered as astrocytic LDs. All the assayed LDs volumes were normalized by the corresponding number of cells. The MAP2 and GFAP volumes were also calculated after proper thresholding in a 3D model of Cellprofiler to measure the neuron-occupied areas. ***CellRox staining***: Since all CellRox signals were located in mAst, their signal volumes were calculated without any separation in a 3D model of Cellprofiler after proper thresholding (20x objective lens). The number of mAst was also quantified to normalize CellRox signals. ***SLC7A11 staining***: The analysis method was the same as for LipidTox staining. Briefly, SLC7A11 signals (20x objective lens) were separated from MAP2 staining. Neuronal and astrocytic SLC7A11 was identified based on MAP2 3D mask in Cell Profiler. mAst cell number was obtained by subtracting MAP2+ neuron count from total cell counts. SLC7A11 volume per mAst was calculated as the total astrocytic SLC7A11 volume divided by mAst cell number.

#### RNA-seq data analysis

Raw FASTQ sequencing reads were trimmed by trim_galore and mapped to a concatenated reference genome of human (GRCh38) and mouse (GRCm39) by using Salmon (v0.11.3). All mapped transcripts (310,359, human and mouse) were further filtered based on the criterion that ≥75% of samples must express a transcript. Filtered human (40,162) and mouse (32,476) transcripts were analyzed for differential expression (DE) between genotypes (TT vs. CC) by using EdgeR (v4.0.16) with quasi-likelihood negative binomial generalized log-linear model. The ratio of human and mouse read counts generated from HISAT2 (v2.1.0) alignment for each sample was used as a correction factor to balance the cellular composition variance of human and mouse cells in iGlut-mAst co-culture. Counts per million (CPM) values of all the filtered transcripts were used for PCA analysis. Transcripts showing significant DE were identified with FDR<0.05, and the top-ranked transcripts were used to represent genes ^109^. Enrichr ^110^ was used to perform GO (Gene Ontology) and Wiki pathway enrichment analyses with a statistical significance cut-off of adjusted p value<0.05.

#### Statistical analyses

Two-tailed unpaired Student’s t-test or Mann-Whitney test (if the data are not normally distributed) were used to test the differences between two groups (TT vs.CC or vehicle vs. treatment). For the test of three groups (AAV-CLU overexpression assay), one-way ANOVA followed by a post hoc Tukey’s test was used to determine significance between groups. The data were analyzed by R 4.1.1 or GraphPad Prism 9. The statistical significance cut-off was 0.05.

